# Can Rights-Based Conditional Cash Transfers Improve Children’s Nutrition at scale? Evidence from India’s Maternity Benefit Program

**DOI:** 10.1101/2025.01.12.25320443

**Authors:** Soumyajit Ray, Suman Chakrabarti, Sumantra Pal, Phuong Hong Nguyen, Samuel Scott, Purnima Menon

## Abstract

This study evaluates the impact of India’s Pradhan Mantri Matru Vandana Yojana (PMMVY), a large-scale conditional cash transfer (CCT) program targeting women during their first birth, on child nutrition. Using National Family Health Survey (NFHS) data from 2005 to 2021, we assess changes in growth for 296,782 children under five years old before and after PMMVY implementation. To address potential biases, we employ a quasi-experimental approach with a Triple Difference analysis, comparing first- to second-born children of CCT and non-CCT mothers. We find that potential exposure to PMMVY is associated with improvements in weight-for-age and height-for-age z-scores. These effects likely operate through increased pregnancy registration, antenatal care, and immunizations. PMMVY is cost-effective, with a short-run benefit-cost ratio of 1.35. This study underscores the importance of CCT programs targeting mothers in enhancing child nutrition in low- and middle-income countries.

## 1 Introduction

Low utilization of primary, preventive health care during pregnancy and early childhood is a key determinant of suboptimal maternal and child health outcomes, particularly in low- and middle-income countries (LMICs) (de Groot et al., 2017). Conditional Cash Transfer (CCT) programs incentivize low-income households to align their behaviour with national social objectives by directly providing cash when they meet specific conditions, such as ensuring children’s school attendance or receive immunizations (Fernald et al., 2008). Some cash transfers are designed to increase the demand for health interventions and enhance uptake of primary health care (Manley et al., 2022). Currently, over 1.3 billion people in 100 countries have access to cash transfers (de Groot et al., 2017; Manley et al., 2022; Manley and Slavchevska, 2019; Owusu-Addo et al., 2018; Richterman et al., 2023). In various contexts, cash transfers have been associated with reductions in the risk of death among young children (Richterman et al., 2023), improved child health and nutrition outcomes among economically disadvantaged families (Galicia et al., 2016), and improved household diet quality (Manley et al., 2022). Viewing cash transfers as strategic investments to enhance nutritional status during a child’s early years holds potential for long-term returns at individual and national levels. For India, investments made for a plausible set of nutritional interventions to reduce stunting are estimated to deliver a 34-fold return on investment in economic benefits (Hoddinott et al., 2013). The potential returns in India are particularly high given the large number of individuals who are positioned to benefit from such programs.

In India, from 2005 to 2016, the Janani Suraksha Yojana (JSY) was the sole nationwide perinatal CCT program, providing coverage to over 10 million pregnant women **(Table S1)** (Carvalho et al., 2014; Lim et al., 2010). However, in January 2017, the Indian Government introduced the Pradhan Mantri Matru Vandana Yojana (PMMVY) (which roughly translates to Prime Minister’s Maternity Benefit Program in English), another CCT program specifically catering to pregnant and lactating women in all districts throughout the country (Government of India, 2022). Every Indian pregnant and lactating woman is entitled to a maternity benefit of a minimum 6,000 Indian rupees (INR) under the National Food Security Act (NFSA) 2013. By capitalizing on the extensive reach and scope of the JSY, the PMMVY is potentially the world’s largest perinatal CCT program in terms of number of individuals reached (Lim et al., 2010). As on March 2024, PMMVY program has enrolled 38.3 million beneficiaries and has made cash transfers to 33.8 million beneficiaries.

While JSY money was conditional on institutional childbirths, offering a cash incentive ranging from INR 1,000 (rural) to INR 1,400 (urban) for women below the poverty line, the PMMVY extends support to all eligible Indian women, granting INR 5,000 (approximately US$ 60 in 2024) (the sum of JSY and PMMVY fulfils the NFSA entitlement) for their first live birth upon fulfilling certain conditions, including pregnancy registration, receiving antenatal care, birth registration, and ensuring the administration of essential vaccinations for newborns (Government of India, 2022). The PMMVY’s design draws inspiration from existing state specific CCT programs, namely the Muthu Lakshmi Scheme in Tamil Nadu and the Mamata Scheme in Odisha (Balasubramanian and Ravindran, 2012; Raghunathan et al., 2017). Although prior studies have demonstrated the positive impact of state-specific schemes on enhancing the odds of receiving of essential health and nutrition interventions (counselling for breastfeeding, antenatal care, and vaccinations), improved food security outcomes, and reducing undernutrition (child stunting and anaemia), there is currently insufficient evidence regarding the effectiveness of the PMMVY on these outcomes (Balasubramanian and Ravindran, 2012; Chakrabarti et al., 2021; Raghunathan et al., 2017).

In this study, we assessed the impact of the PMMVY on children’s nutrition outcomes. Our descriptive analyses revealed that the coverage of PMMVY in its first four years falls below that of successful predecessor CCTs like the JSY. Nonetheless, the program has achieved considerable scale in targeting firstborn children, albeit with some instances of second born children also being covered. Through the analysis of three rounds of India’s National Family Health Surveys (NFHS) (2005-06, 2015-16 and 2019-21), we investigated changes in key child anthropometric outcomes among over 300,000 surveyed children in India, comparing periods before and after the implementation of PMMVY across appropriate comparison groups.

One of the primary challenges in evaluating whether cash transfers improve outcomes is identifying a plausible counterfactual: what would a child’s nutritional status have been in the absence of receiving transfers? PMMVY was launched nationwide, during a period of increasing spending on healthcare, thus, any improvements in child nutrition after PMMVY could reflect broader trends and not be caused by PMMVY. In such scenarios, the most widely used method for isolating causal effects of transfer programs is adopting a difference-in-differences (DID) design using panel data. For PMMVY, we use pooled data from NFHS-4 (2015-16) and NFHS-5 (2019-21) on infants and young children. Children aged 0-5 years born before 2017 were untreated cohorts (pre-PMMVY) and those born after 2017 were treated cohorts (post-PMMVY). The second requirement for DID is identifying an appropriate control group. Since the NFHS did not directly ask women if they received PMMVY benefits, the next best option is to compare women who received any perinatal CCT to women who did not. Since PMMVY registration is implemented by the same frontline workers who register beneficiaries for the JSY and other pre-existing perinatal CCTs (already nationally implemented for over 10 years), women who opt for existing CCTs are also likely to opt for the PMMVY. Comparing changes in outcomes in the CCT group versus changes in outcomes in the non-CCT groups would provide an intent to treat DID estimate for CCT beneficiaries. However, this DID estimate is prone to potential biases because CCT and non-CCT women may have large differences in observed and unobserved characteristics, which may result in selection bias, even if the pre-intervention parallel trends assumption was satisfied. Moreover, the CCT group contains children of higher birth orders, who were not entitled to receive PMMVY money, further skewing the DID estimate downwards, and not isolating the impact of PMMVY.

To account for both sources of bias, we add a third axis of comparison; comparing the DID between firstborns (treated children) to second born children (non-treated) (Olden and Møen, 2022). The resulting Triple Difference (TD) estimate is credible because unobservable characteristics of first and second born children within the CCT and non-CCT groups would be similar (Aronow and Miller, 2019). For example, within the CCT group, mothers of first and second born children would likely have similar wealth, height, and education. Further, omitted variables that distinguish first and second born children in India would likely be similar across the CCT and non-CCT groups. For example, any knowledge of best practices from firstborns would get transmitted during the care of second born children, and this transmission is expected to be similar across CCT and non-CCT groups.

Employing TD analyses, we showed that exposure to the PMMVY was associated with modest improvements in WAZ (0.05 SD) and HAZ (0.08 SD) among firstborn CCT children. Using an indirect placebo test with a fake treatment, the parallel trends assumption was not rejected for both outcomes in the TD model. To investigate potential mechanisms behind the positive effect on anthropometric outcomes, we estimated the effect of the program on conditionalities. PMMVY increased the odds of pregnancy registration, antenatal care, and immunizations, by 12%, 6% and 10%, respectively. These findings suggest PMMVY likely improves child anthropometric outcomes though program conditionalities. Exploring heterogeneities in coefficients, we found that PMMVY impacts were larger and significant when the program was delivered by the health ministry compared to other entities, suggesting that prior experience with national transfers (JSY) may have played a role for program success. Moreover, we found that program impacts for HAZ were twice as large for children from poor households (0.10 SD) compared to non-poor households (0.05 SD). Impact coefficients for HAZ also varied by sex with significant impacts for males (0.14 SD) but not for females (0.06 SD), suggesting that higher son preference among beneficiary mothers may have resulted in higher allocation of financial resources towards boys.

Next, we examined performed a benefit-cost analysis of PMMVY for its three-year impact. For this, we fit a state-level birth cohort model for firstborns born between 2017 to 2020. We use program expenditure data from the Indian parliament as the explanatory variable to test if differences in per head program spending across states and birth cohorts predict an increase in anthropometry. Here we find that INR 1,000 per head higher program expenditure is associated with a 1.27 percentage point (pp) reduction in underweight and 1.38 pp reduction in stunting, among first born CCT children, compared to non-CCT children. Building upon this expenditure-outcome relationship, we subsequently estimate the economic returns of the program. Economic analyses suggest that PMMVY delivered substantial health benefits, with a short-run benefit-cost ratio of 1.35, indicating cost-effectiveness.

Current implementation challenges of the PMMVY program include lengthy documentation for enrolment, linking bank accounts with social security numbers, and securing cooperation from frontline workers and child development project officers. These obstacles hinder access to the program, especially for women with limited education. Payment delays further limit the timely utilization of funds. Despite these challenges, our study shows that PMMVY is cost-effective and implemented at scale, benefiting from India’s past experience with cash transfer programs. The recent (2022) extension of PMMVY benefits to second-born girl children indicates broader future coverage. Our findings suggests that CCT programs like PMMVY hold promise for improving child health and well-being in India and beyond.

The subsequent section of this paper is organized as follows: Section 2 discusses the history of maternal cash transfer programs in India, with a focus on the PMMVY program. Section 3 outlines the data sources used in the study. Section 4 describes the empirical strategy used to assess the impact of the PMMVY program. Then, Section 5 present our key findings, and Section 6 demonstrates the heterogeneous impact of the program. Section 7 provides a benefit-cost analysis for the program. Finally, Section 8 concludes by presenting the paper’s strengths and limitations, while underscoring implementation challenges.

## 2 Background

### 2.1 Maternal Cash Transfer Programs in India

India has a long history of designing and executing large-scale social protection programs for the poor and marginalized communities (Drèze and Khera, 2017; Kapur and Nangia, 2015; Sen and Rajasekhar, 2012). Examples of such programs include housing programs (Pradhan Mantri Gramin Awas Yojana) for the rural poor and workfare programs (Mahatma Gandhi National Rural Employment Guarantee Act), among others. The national government implements various food and nutrition programs such as the Public Distribution System for households below the poverty line, Integrated Child Development Services for children between 0 and 6 years, and the Mid-Day Meal program for children in the upper primary classes (Chakrabarti et al., 2021, 2019; Kishore and Chakrabarti, 2015).

Governments at both the national and regional levels have implemented programs exclusively targeting pregnant women (Bhatia et al., 2006; Godha and Hotchkiss, 2022). These programs provide conditional or unconditional cash directly to the beneficiaries, vouchers to pregnant women for seeking healthcare services for free, or a combination of both (Chowdhry, 2013). Since the 1980s, maternal cash-only transfer programs have been implemented, as shown **Figure 1 (Table S1)**. Tamil Nadu, a southern state in India, launched the first maternal cash transfer program called the Dr. Muthulakshmi Reddy Maternity Benefit Scheme in 1987, aimed at reducing infant and maternal mortality rates. In 1995, the Government of India launched its own maternal cash transfer scheme, the National Maternal Benefit Scheme, covering women from poor households for two live births. This program was later revamped in 2005 and launched as JSY to promote institutional delivery, especially in rural areas (Bhatia et al., 2006).

**Figure 1.**
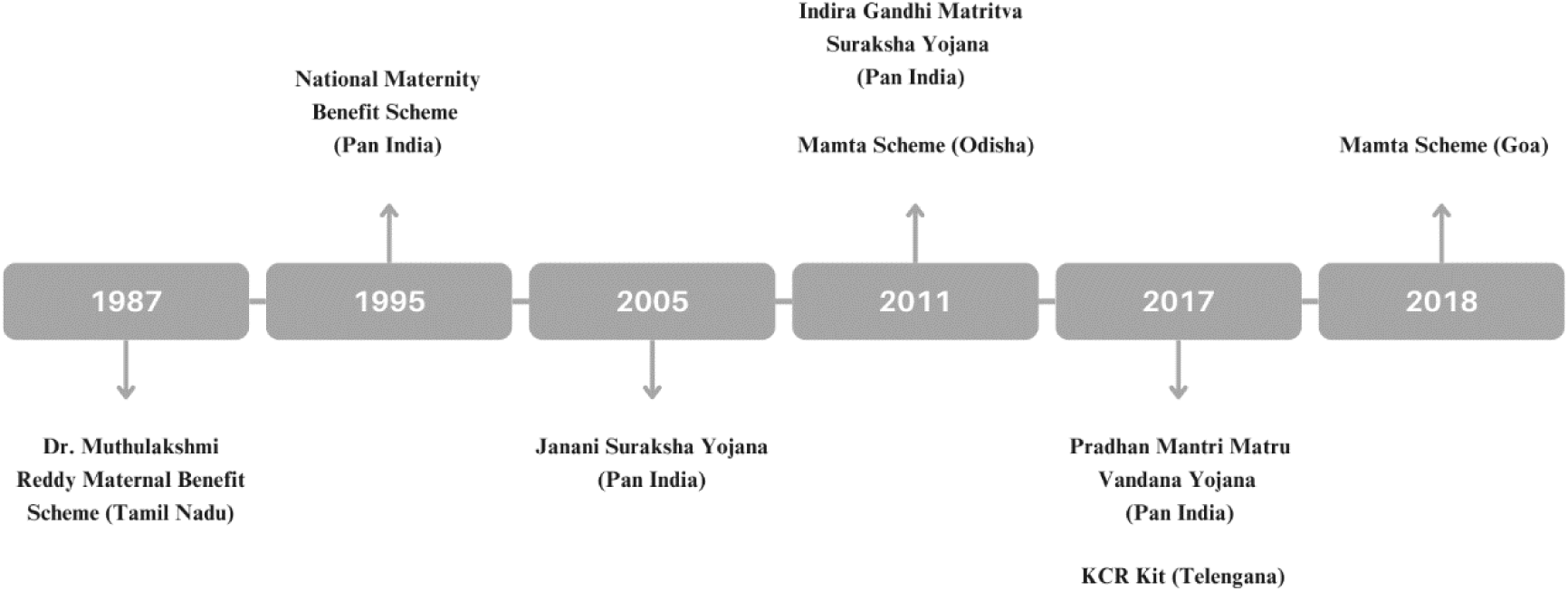
Landscape of India’s maternal cash transfer programs, 1987-2021. Note: Telangana and Tamil Nadu are situated in southern India, while Odisha is located in the eastern part, and Goa is positioned in western India. The Mamta Scheme refers to a scheme for mothers. KCR Kit in Telangana is named after Chief Minister K. Chandrasekhar Rao, under whose tenure the program was launched.

Success attained by JSY in increasing antenatal care and in-facility births, along with a reduction in prenatal and neonatal births, inspired additional maternal cash transfer programs in the following decade (Lim et al., 2010). The Indian government piloted the Indira Gandhi Matritva Sahyog Yojana (IGMSY) program in 52 of the 640 districts between 2011 and 2016, providing INR 4,000 for the first two live births (Haaren and Klonner, 2021; Sinha et al., 2016). During the same period, the state government of Odisha launched the Mamata Scheme in Odisha, disbursing a total of INR 5,000 to provide partial wage compensation to pregnant and nursing women for the first two live births (Chakrabarti et al., 2021; Patwardhan, 2023).

In January 2017, the Government of India revamped the IGMSY program to PMMVY, catering to pregnant and lactating women throughout the country for the first live birth. The government in the state of Telangana launched its own conditional maternal cash transfer program around the same time for economically weaker sections of society. In the subsequent year, the Government of the state of Goa launched its own Mamata scheme providing INR 10,000 to women for delivering a girl child at a registered medical facility.

### 2.2 The PMMVY Program

Under PMMVY, a sum of INR 5,000 is transferred in three instalments to eligible beneficiaries for their first live birth upon fulfilling certain conditionalities. In theory, a combination of PMMVY and JSY would provide a beneficiary INR 6,000 (INR 6,400) in urban (rural) areas. The objectives of PMMVY are two pronged: firstly, to offer partial income compensation to allow a pregnant/lactating woman appropriate rest before and after the delivery of her first child; and secondly to enhance pregnant and lactating women’s health and nutrition seeking practices and behaviour (Government of India, 2022).

PMMVY is a collaborative effort between both the national and state governments, with each sharing the financial cost of the implementation. In Union Territories, which are designated as special administrative regions governed by the central government, the entire cost of the program is borne by central government. In hilly areas and states with special status, the central government provides 90% of the funding, with the remaining 10% contributed by the state governments. Across the rest of the country, the central government funds 60% of the program, with state governments covering the remaining 40%. The Ministry of Women and Child Development (MWCD) oversees the program’s implementation at the national level, while individual state governments can house the program at the MWCD, health department, or social welfare and justice departments.

Details on eligibility, supporting documents required for processing the payments, and timing of payments are provided in **Table S2**. In the period we cover for the current analysis, payments were made in three instalments. For their first pregnancy, women receive the first instalment of INR 1,000 upon completion of pregnancy registration within 150 days of pregnancy. The second instalment of INR 2,000 is after 180 days of pregnancy conditional and upon women completing at least one antenatal care check-up. The third instalment of INR 2,000 is paid upon completing childbirth registration and the first cycle of BCG, OPV, DPT and Hepatitis B vaccinations for the firstborn child.

## 3 Data, program coverage, outcomes, and covariates

### 3.1 Data Sources

For impact assessments, we used mother-child and household-level data from three rounds of the Indian National Family Health Surveys (NFHS, equivalent to Demographic Health Surveys in other countries) in 2005-06 (NFHS-3), 2015-2016 (NFHS-4) and 2019-2021 (NFHS-5). We refer to these surveys as the 2005, 2015, and 2020 rounds from here on. These cross-sectional surveys follow a systematic, multi-stage stratified sampling design, covering all states/union territories in India. While the 2005 round is representative at the country and state level, the 2015 and 2020 rounds are representative at country, state and district levels. Given that PMMVY was launched in 2017, the 2005 round allows us to examine pre-intervention secular trends, the 2015 round serves as the pre-intervention baseline and the 2020 round provides post intervention period estimates, facilitating a pre-post comparison. We use data on the youngest child for every mother in the sample because data on perinatal cash transfers is only available for these children. We exclude Tamil Nadu, Puducherry, Odisha, and Telangana from the analyses because they already or simultaneously implemented state-wide perinatal cash transfers (Chakrabarti et al., 2021; Patwardhan, 2023; Raghunathan et al., 2017). We supplement NFHS data with cash disbursement data on PMMVY from the data bank of India’s parliament (Ministry of Women and Child Development, 2021) along with population data from the Health Management Information System and the Indian census to conduct a benefit-cost analysis of PMMVY.

The pooled NFHS-4 and NFHS-5 samples comprised N=367,741 children under five years old. After excluding children without valid anthropometric measurements (N=38,174), those from Tamil Nadu, Puducherry, Odisha, and Telangana (N=32,785) and third and higher birth order children (N=100,791), the final analysis sample was N=296,782 children under five years old **(Figure S1)**. For all our analyses, we exclude all third-born and subsequent children because previous literature suggests that high birth order children are more likely to experience growth faltering in India and are systematically different from their older siblings (Jayachandran and Pande, 2017; Spears et al., 2019). **Figure S2** shows that third and higher birth orders were very different from first and second born children in trends and outcome levels. The remaining sections focus on first and second born children for descriptives, identification strategy for our empirical model, and robustness checks.

### 3.2 Disbursement and coverage of PMMVY

We estimate coverage of PMMVY in two ways. First, we use the total number of women who registered for PMMVY between 2017 and 2021. To convert absolute numbers to percentages, we use total live births in this period from Health Management Information System and the birth order distribution of Indian children from the Census to estimate an upper bound for PMMVY coverage. Per Census and HMIS data, out of the 43.56 million firstborn children, 50.8% were covered by PMMVY between 2017 and 2020 on average. **Figure 2** shows the trend in coverage of PMMVY since its rollout in 2017, with 63% of eligible beneficiaries being reached in 2019–20. However, the per-capita disbursement as a proportion of total eligible beneficiaries was low, with the highest being INR 2,953 (approximately US$ 36) per capita spending on the eligible population for 2019-20. Among beneficiaries who received PMMVY funds, disbursements ranged between INR 1,106 to INR 4,690, on average.

**Figure 2.**
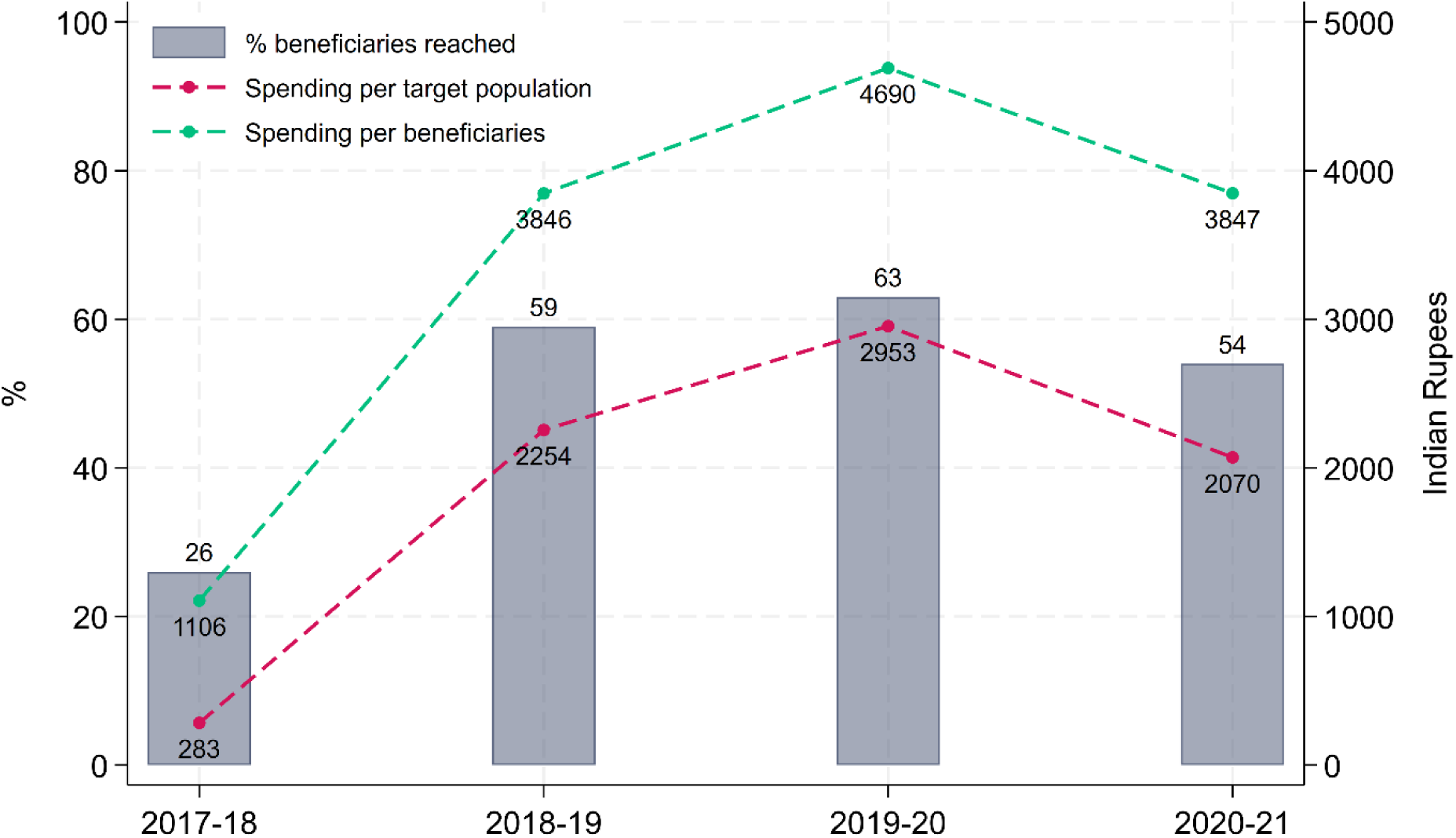
Resource allocation and program utilization for PMMVY between 2017 and 2021. Notes: PMMVY refers to Pradhan Mantri Matru Vandana Yojana. The coverage and expenditure figures are based on data from the Health Management Information System, the Census Sample Registration System, and responses filed by the Ministry of Women and Child Development in the Indian parliament.

Second, the 2015 and 2020 NFHS rounds also provide data on women who received money from the JSY or any other perinatal cash transfer; along with the amount they received from JSY. It does not, however, ask any direct question about PMMVY. In the absence of a specific PMMVY variable, we calculated the percentage of women who reported receiving money from any perinatal cash transfer and those who reported receiving INR 5,000 or more as a proxy for PMMVY coverage. However, the proportion of mothers receiving INR 5,000 or more, is likely an underestimate of PMMVY coverage because women familiar with JSY would report accurate amounts (around INR 1,400), whereas women unfamiliar with JSY may report higher amounts if they benefited from other transfers. Therefore, the ‘proxy’ PMMVY coverage variable from NFHS is only used for descriptive analyses and to validate our identification strategy. Per NFHS data, in 2015, mothers of 34.5% of firstborns and 35.8% of second born children received any perinatal CCT **(Figure 3, Panel A)**. Coverage among firstborns (36.5%) increased by 2020 but decreased for the second born children (32.5%). Between 2015 and 2020, among JSY beneficiaries, mothers who received at least INR 5,000 increased from 1.3% to 22.5% for firstborns **(Figure 3, Panel A)**. Together, these statistics suggest that PMMVY was rolled out between 2015-16 and 2019-21, and firstborns received the most benefits. However, 5.6% of mothers did report receiving INR 5,000 for their second born child **(Figure 3, Panel B)**. This may be attributed to other cash transfer programs, instances where the firstborn child may have died, or mistargeting of PMMVY.

**Figure 3.**
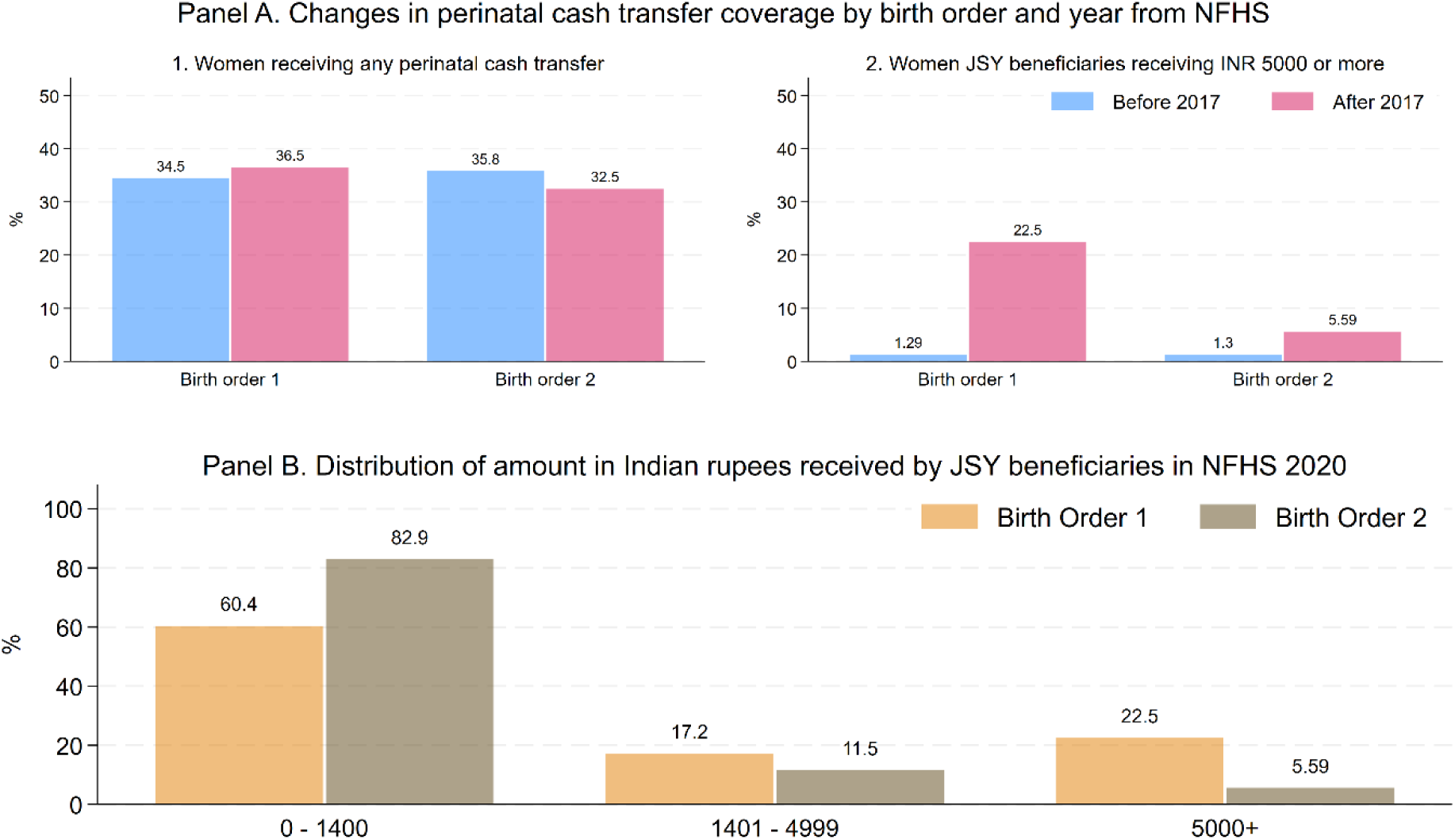
Coverage of perinatal cash transfer programs in India based on the NFHS 4 and 5. Notes: States with existing maternal benefits programs, such as Tamil Nadu, Odisha, and Puducherry, were dropped from the NFHS sample to isolate the PMMVY beneficiaries. Indicators on cash transfer and amount of cash received are based on the question asked for JSY in the NFHS.

### 3.3 Outcomes

Child height and weight were measured by well-trained researchers using SECA-874U digital scales, SECA-213 stadiometers and SECA-417 infantometers (International Institute for Population Sciences, 2022). We use age (in months), height (in centimetres), weight (kg) and the WHO age-sex growth standards to calculate the z-scores for children aged 0 to 5 years (de Onis, 2006; Vidmar et al., 2013). The outcomes of interest are height-for-age z-score (HAZ), weight-for-age z-score (WAZ), stunting (HAZ<-2 SD) and child underweight (WAZ<-2 SD). While WAZ and HAZ respectively measure child weight and linear growth on a continuous scale relative to healthy children globally, underweight and stunting indicate if a child is undernourished with respect to global standards. All four outcomes are globally recognized as markers of undernutrition among children and have been prioritized under the sustainable development goals (Fullman et al., 2017).

### 3.4 Covariates

Child-level covariates include age (months), sex (male/female). Mother-level covariates include height (cm), body mass index (kg/m^2^), age (years), and education (years). Household-level covariates include health insurance (binary), family size (number), religion (Hindu, Muslim, Christian), caste (disadvantaged, tribal) and wealth quintile (ranging from 1-5). A wealth index was constructed with a principal component analysis of household’s characteristics including source of drinking water, type of toilet facilities, type of flooring, exterior wall material, type of roofing, cooking fuel, electricity, home ownership, domestic helper, number of household members per sleeping room, ownership of a bank or post office account and having a mattress, pressure cooker, chair, cot/bed, table, electric fan, radio/transistor, black-and-white television, color television, sewing machine, mobile phone, any other telephone, computer, refrigerator, watch or clock, bicycle, motorcycle or scooter, an animal-drawn cart, car, water pump, thresher, tractor and livestock (cows,camels, goats, horse, chicken, pigs) (Filmer and Pritchett, 2001). The index was constructed after pooling the NFHS rounds to obtain a consistent measure of asset poverty over time. In other words, within wealth quintiles, households have the same set of assets and amenities across NFHS rounds. Among the covariates included, maternal height, education, BMI, and household wealth, in that order, are among the strongest predictors of child undernutrition in India (Li et al., 2020). The remaining variables are standard socio-demographic controls that account for residual variation and improve precision.

## 4 Empirical Strategy and Identification of the Effects of Cash Transfers

Ideally, random assignment of women to PMMVY would allow causal estimation of the average effect of the intervention. However, in the absence of a randomized assignment of individuals to treatment (i.e., exposure to PMMVY), we treated the timing and targeting of PMMVY as a natural (or quasi) experiment. For PMMVY, there may be unobservable child-specific factors (e.g., location, ingrained dietary habits, etc.) that are associated with receiving a cash benefit and child undernutrition (de Groot et al., 2017; Fernald et al., 2008). Such factors, however, are likely to be relatively invariant over the short-term (Shadish et al., 2002). There may also be time-varying factors that could bias estimates, such as the national implementation of child health and nutrition programs. The preferred method of controlling for both issues is to use longitudinal data and estimate difference-in-differences (DID) models (Angrist and Pischke, 2009; Shadish et al., 2002). The DID estimation strategy requires that (1) outcome data be available before and after the intervention and (2) treatment and control (comparison) groups can be clearly distinguished. Here, examination of temporal changes within the treated group (between 2016 and 2021) controls for factors that don’t change in the short term. Accounting for change in outcomes within the comparison group acts as a counterfactual estimate for what would have happened in the absence of PMMVY and controls for factors that may change over time but are common to both groups. In other words, by looking at the changes in the treated group, while taking into account changes in the comparison group before and after the intervention, we can obtain the average treatment effect estimates (Angrist and Pischke, 2009). However, the PMMVY’s unique features offer an opportunity to compare multiple groups in a Triple Difference (TD) framework to further account for remaining biases (Olden and Møen, 2022). Such biases may stem from time varying factors that are group specific, for instance rates of economic growth among low SES households (that are more likely take up PMMVY) may have trended differently compared to high SES households. The triple difference estimator can be computed as the difference between two difference-in-differences estimators with two comparison groups.

### 4.1 Comparison group 1: perinatal cash transfer non-beneficiaries

Finding an appropriate comparison group for PMMVY requires identifying children who were not eligible to receive PMMVY money, before and after PMMVY was implemented. As mentioned earlier, there is no variable that directly identifies PMMVY beneficiaries in the NFHS. However, the PMMVY selection mechanism is likely to resemble processes present in other pro-poor perinatal cash transfers like the JSY for which variables are available in NFHS (Carvalho et al., 2014; Lim et al., 2010). Even though PMMVY is available to mothers from all households, beneficiaries are likely to belong to lower SES groups because the INR 5,000 entitlement would not likely incentivise upper SES households. In other words, mothers who take-up the JSY (already nationally implemented for over 10 years) are also likely to opt in for the PMMVY. Thus, as our first comparison group, we compared any perinatal CCT beneficiaries to non-beneficiaries.

Yet, this comparison group is likely to suffer from selection bias stemming from the demand side; PMMVY non-beneficiaries may make health-specific demand choices due to unobserved factors that are systematically different from beneficiaries. Even if pre-intervention parallel trends would hold, the groups may trend differently overtime due to large differences in their covariates. Further, in this comparison, it would be difficult to isolate the effects of PMMVY money from other perinatal CCTs. Finally, even the CCT group contains second born children, who were not eligible for PMMVY. Taking these concerns together, any DID estimates comparing CCT to non-CCT children would be likely be biased.

### 4.2 Comparison group 2: second born children

To account for sources of bias mentioned above, we add another axis of comparison, comparing the CCT versus non-CCT DID between firstborns (treated children) to second born children (non-treated) (Olden and Møen, 2022). The resulting TD estimate is more credible because unobservable characteristics of first and second born children within the CCT and non-CCT groups would likely be similar (Aronow and Miller, 2019). In other words, adding another control group improves exchangeability of participants in the sample, making them similar on unobservable variables that may confound results (Aronow and Miller, 2019). Exchangeability refers to the assumption that, after conditioning on observed variables, the treated and control groups have similar potential outcomes, enabling estimation of causal effects from observational data. For example, first and second born children among CCT and non-CCT mothers would likely have similar wealth, maternal height, BMI, and education, constituting the strongest known confounders for child undernutrition (Table 1). Further, omitted variables that distinguish first and second born children in India would likely be similar across the CCT and non-CCT groups. For example, any knowledge of best practices from firstborns would get transmitted during the care of second born children, but this transmission is likely similar across CCT and non-CCT groups. The validity of TD on balancing observed covariates is discussed in the results section **(Table 1)**.

**Table 1.** Summary statistics of covariates among comparison groups in preintervention period.

|  | Conditional cash transfer (CCT) group |  |  |  |  | Non-Conditional cash transfer (non-CCT) group |  |  |  |  |  |
| --- | --- | --- | --- | --- | --- | --- | --- | --- | --- | --- | --- |
|  | Birth order 1 |  | Birth order 2 |  | CCT difference<br>BO1-BO2 | Birth order 1 |  | Birth order 2 |  | non-CCT difference<br>BO1-BO2 | Triple difference |
|  | Mean/% | 95%CI | Mean/% | 95%CI | Mean/% | Mean/% | 95%CI | Mean/% | 95%CI | Mean/% | Mean/% |
| Mother's height, cm | 151.7 | [151.6,151.8] | 151.9 | [151.8,152.0] | -0.2 | 152.4 | [152.3,152.5] | 152.2 | [152.1,152.3] | 0.2 | -0.4 |
| Mother's BMI, kg/m<sup>2</sup> | 20.6 | [20.6,20.7] | 20.9 | [20.8,20.9] | -0.3 | 21.6 | [21.5,21.6] | 21.7 | [21.7,21.8] | -0.1 | -0.2 |
| Mother's age, year | 23.4 | [23.4,23.5] | 26.1 | [26.0,26.1] | -2.7 | 24.1 | [24.1,24.2] | 26.6 | [26.5,26.7] | -2.5 | -0.2 |
| Mother's education, year | 7.8 | [7.7,7.9] | 6.6 | [6.5,6.6] | 1.2 | 9.2 | [9.1,9.3] | 7.7 | [7.6,7.8] | 1.5 | -0.3 |
| Socio-economic status score, 1-5 | 2.5 | [2.5,2.6] | 2.5 | [2.5,2.5] | 0 | 3.2 | [3.2,3.2] | 3.0 | [3.0,3.1] | 0.2 | -0.2 |
| Health insurance, % | 22.8 | [21.9,23.7] | 22.7 | [21.8,23.6] | 0.1 | 38.3 | [21.2,22.6] | 20.7 | [20.0,21.4] | 1.2 | -1.1 |
| Family size, # | 5.7 | [5.7,5.8] | 6.0 | [6.0,6.1] | -0.3 | 21.9 | [5.7,5.8] | 6.2 | [6.1,6.2] | -0.4 | 0.1 |
| Hindu, % | 82.0 | [81.2,82.9] | 83.4 | [82.6,84.1] | -1.4 | 5.8 | [75.8,77.3] | 77.5 | [76.8,78.2] | -0.9 | -0.5 |
| Muslim, % | 13.4 | [12.6,14.1] | 12.2 | [11.6,12.8] | 1.2 | 76.6 | [15.8,17.1] | 16.4 | [15.7,17.0] | 0 | 1.2 |
| Christian, % | 1.6 | [1.3,1.8] | 1.6 | [1.3,1.9] | 0 | 16.4 | [1.9,2.4] | 2.0 | [1.7,2.2] | 0.2 | -0.2 |
| Scheduled caste, % | 23.6 | [22.7,24.5] | 23.3 | [22.4,24.2] | 0.3 | 2.2 | [17.6,19.0] | 18.6 | [17.9,19.3] | -0.3 | 0.6 |
| Scheduled tribe, % | 12.8 | [12.1,13.4] | 11.6 | [10.9,12.2] | 1.2 | 18.3 | [7.6,8.4] | 8.7 | [8.3,9.1] | -0.7 | 1.9 |
| Child age, months | 23.1 | [22.8,23.4] | 26.2 | [25.8,26.5] | -3.1 | 8.0 | [22.4,23.0] | 25.9 | [25.6,26.2] | -3.2 | 0.1 |
| Child sex male, % | 51.6 | [50.6,52.7] | 53.9 | [52.9,54.9] | -2.3 | 22.7 | [53.4,55.1] | 54.7 | [53.9,55.5] | -0.5 | -1.8 |
| Observations | 17819 |  | 18606 |  |  | 31386 |  | 31506 |  |  |  |

Note: Summary statistics are generated using the NFHS 4 data. The sample is limited to only the first and second-born children under 5 years with non-missing anthropometric measurements. Socio-economic status scores are constructed based on principle component analysis of household assets. BMI refers to Body-Mass Index.

### 4.3 Triple difference estimating model

Since this model has two pre-post comparison groups, (1) perinatal CCT versus non-CCT children and (2) first versus second born children, the TD model is the unbiased difference between two DID models that may have been separately biased. In other words, biases in comparing first and second born CCT children are differenced out by using the difference of first and second born non-CCT children as a control. Additionally, biases in comparing CCT and non-CCT firstborns are differenced out by using the difference of CCT and non-CCT second born children as a control. The intuition is that the difference between two biased DID estimators will be unbiased if the bias is the same in both estimators (Olden and Møen, 2022). The effects estimated by equation 1 are intent-to-treat (ITT) estimates because our TD strategy models potential exposure of mothers who are likely to opt for the PMMVY. Our approach does not exclusively identify children who were direct beneficiaries of the PMMVY. ITT is a policy-relevant parameter for an ex-post analysis of the effects of a large policy on the entire target population (Angrist et al., 1996).

Our primary empirical strategy relies on pooling data from NFHS-4 and NFHS-5. To estimate the average treatment effect for PMMVY, we fit a triple difference model with *equation 1* for child *i*, born to mother *m*, from household *h* in community *c* in year of survey *t*:

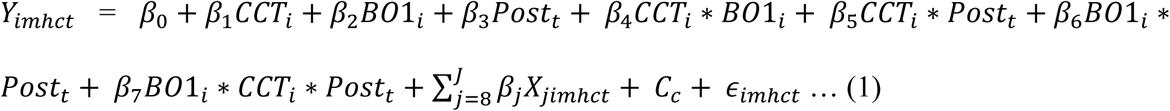

where *Y*_*imhdt*_ is a continuous outcome (HAZ or WAZ). *CCT*_*i*_ is a treatment dummy that takes value 0 for non-CCT firstborn children, i.e. those born to mothers who reported that they did not receive any perinatal cash transfer, and 1 for CCT firstborn children. *BO*1_*i*_ is a treatment dummy that takes value 0 for second born children and 1 for firstborns. *Post*_*t*_ is a time dummy that takes value 0 for data for children born before 2017 and 1 for children born after 2017. *C*_*c*_ are community fixed effects which control for all neighbourhood or village specific time-invariant (in the short run) confounders such as climate, disease burden, and urbanicity, among others. *X*_*jimhdt*_ represents the vector of covariates included at the individual and household level (*β*_*j*_ indicates specific coefficient corresponding to each covariate within *X*_*jimhdt*_). *ϵ*_*imhdt*_ is an error term that represents residual variation. Standard error estimates are clustered at the community-level to account for intra-community correlations.

*β*_0_ is the pre2017 prevalence among non-CCT second born children. *β*_1_ is the mean difference in outcomes in CCT second born children compared to non-CCT second born children before 2017.

*β*_2_ is the mean difference in outcomes for non-CCT firstborns compared to non-CCT second born children before 2017. *β*_3_ is the change in outcomes comparing non-CCT second born children after 2017 to before 2017. *β*_4_ is the mean difference in outcomes before 2017 comparing the difference of CCT firstborns to non-CCT firstborns to the difference of CCT second born children to non-CCT second born children. *β*_5_ is the mean difference in change (before 2017 to after 2017) in outcomes for CCT second born children above and beyond the difference in non-CCT second born children. *β*_6_ is the mean difference in change (before 2017 to after 2017) in outcomes for non-CCT firstborns compared to the change for non-CCT second born children. Finally, *β*_7_ is the mean difference in change (before 2017 to after 2017) in outcomes for CCT firstborns compared to non-CCT firstborns compared to the changes for CCT second born children compared to non-CCT second born children. *β*_7_ is the TD estimator and parameter of interest for our analysis (Muralidharan and Prakash, 2017).

### 4.4 Parallel trends assumption for triple difference models

Like DID models, the underlying assumption for TD models is that the relative outcome of second- and first-born children in the CCT group trend in the same way as the relative outcome of second- and first-born children in the non-CCT group, in the absence of treatment (Olden and Møen, 2022). We test this assumption with a placebo regression using equation 1 using only NFHS4 data. The key difference in the placebo regression is that *Post*_*t*_ is a time dummy that takes value 0 for birth years before 2014 and 1 for 2014, 2015 and 2016. We omit the vector of covariates *X*_*jimhct*_ and *C*_*c*_. Since PMMVY did not exist for cohorts born between 2014-2016 this model measures the impact of a fake treatment on firstborns born between 2014 and 2016 (just before PMMVY). In this test, *β*_7_ should not be significant and should be of a small order for evidence of parallel trends. In other words, we assume that difference of outcomes would have trended similarly for the two groups in the absence of the intervention of interest (Olden and Møen, 2022).

### 4.5 Randomization inference test for triple difference models

To rule out the possibility that our primary results were obtained by statistical chance, we estimated coefficients using Fisher’s exact test using randomization inference. Here, we randomly assigned the three-way interaction term from our main model and ran 1000 iterations using the “RITEST” Stata module (Heß, 2017). The randomization inference procedure tests whether our main result was obtained by pure chance over 1000 randomized treatment assignments. Lower p values in this test indicate the probability that our main results were obtained by pure chance.

## 5 Main results

### 5.1 Distribution of covariates within treatment groups

Table 1 shows summary statistics comparing covariates across the different comparison groups in the study at baseline. As expected, among CCT beneficiaries, families were poorer, on average, and had six family members living in a household. Mothers of firstborns were 2.7 years younger and had 1.2 years more education than those of second born children. However, these differences were largely eliminated after comparing first and second born children within the CCT (1.2 years) and non-CCT groups (1.5 years). Subtracting estimates for firstborns from second born children, largely eliminated differences in most covariates across the CCT and non-CCT groups, lending credibility to the TD design. However, there were 2.3% fewer male firstborn children in the CCT group, likely because parents may try for a male child if the firstborn was female (Pande and Malhotra, 2006). Thus, son preference presents one potential avenue of bias for the TD models, which we later explore in heterogeneity analyses.

### 5.2 Trends in child anthropometry and changes after PMMVY

We observed similar trends in outcomes between 2005 and 2015 for first and second born children in the sub-sample of perinatal CCT beneficiaries **(Figure 4, Panel A).** Trends appear to deviate after 2015, with firstborns showing faster improvements than second born children in this period. Similarly, trends pre 2015 were similar for CCT and non-CCT beneficiaries among firstborn children for both anthropometric outcomes **(Figure 4, Panel B)**. However, for HAZ, trends deviate after 2015 for CCT beneficiaries.

**Figure 4.**
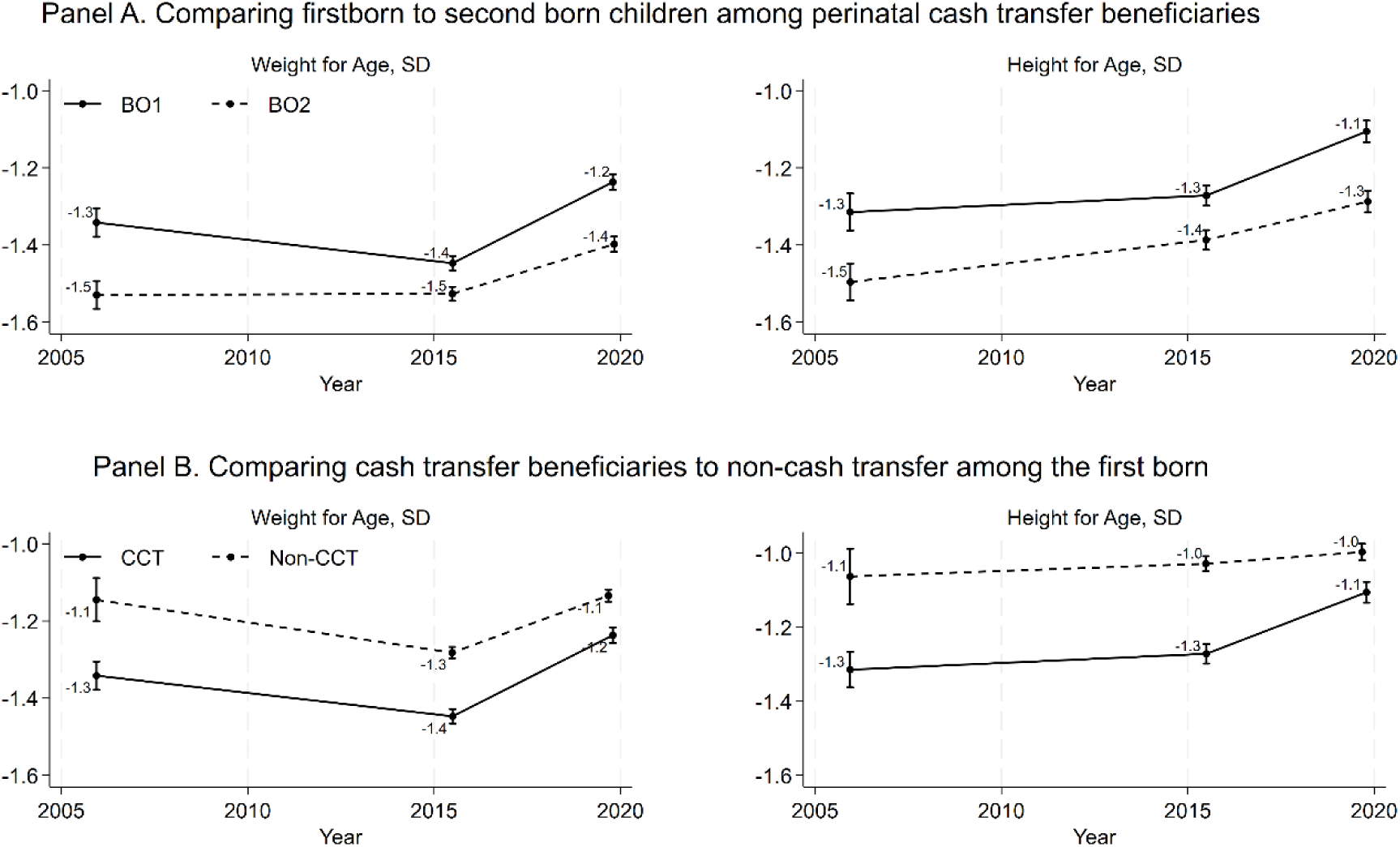
Trends in anthropometric outcomes among Indian children 0-5 years, 2005-2021. Notes: Plots represent the pooled data from NFHS 3, 4, and 5. Weight for age and Height for age z-scores calculated for birth order 1 (BO1) and birth order 2 (BO2), along with those who received conditional cash transfer (CCT) and those who didn’t (non-CCT). Because no national perinatal CCT programs existed in 2005, we predicted the probability of receiving a perinatal CCTs for mothers in 2005 using a logit regression model using data from 2015 and 2020. We use the full set of child, mother, and household level covariates and state fixed effects as predictors. A probability threshold (>0.2) where there was maximum improvement in accuracy of correctly predicting a perinatal CCT beneficiary in 2015 and 2020 was used to classify potential CCT beneficiaries in 2005.

Figure 5 **(also Table S3)** shows the TD impact coefficients (*β*_7_) estimated using equation 1. In the unadjusted TD model, PMMVY was significantly associated with higher WAZ (0.11 SD, p<0.001) and HAZ (0.11 SD, p<0.001). In the model with community fixed effects which accounted for all unobserved time invariant community level confounders, PMMVY was significantly associated with higher WAZ (0.08 SD, p<0.001) and HAZ (0.12 SD, p<0.001). In the fully adjusted model, which included community fixed effects and the full set of covariates, PMMVY was significantly associated with higher WAZ (0.05 SD, p<0.001) and HAZ (0.08 SD, p<0.001). The direction and magnitude of coefficients on HAZ and WAZ for PMMVY estimated in the TD model are in line with other studies evaluating maternal CCTs such as the 2005 JSY, IGMSY, or Mamta scheme in Odisha in the Indian context (Chakrabarti et al., 2021; Ghosh and Kochar, 2018; Kekre and Mahajan, 2023; Patwardhan, 2023; von Haaren and Klonner, 2021). For example, Patwardhan (2023) estimated that WAZ improved by 0.16 SD after the implementation of Mamata, and Kekre and Mahajan (2023) estimated a WAZ impact of 0.14 SD. Similarly, whereas, studies on Mamata and IGMSY reported modest and insignificant effects on HAZ, Chakrabarti et al (2019) found 10% lower odds of stunting among CCT beneficiaries in Odisha. Moreover, global systematic reviews suggest that, on average, CCTs may improve HAZ by 0.024 SD, indicating a marginally higher impact of PMMVY for this outcome (Fernald et al., 2012; Manley et al., 2022). Unlike most global CCTs which do not report significant impacts on WAZ, our models indicate that targeted CCTs can significantly improve this outcome.

**Figure 5.**
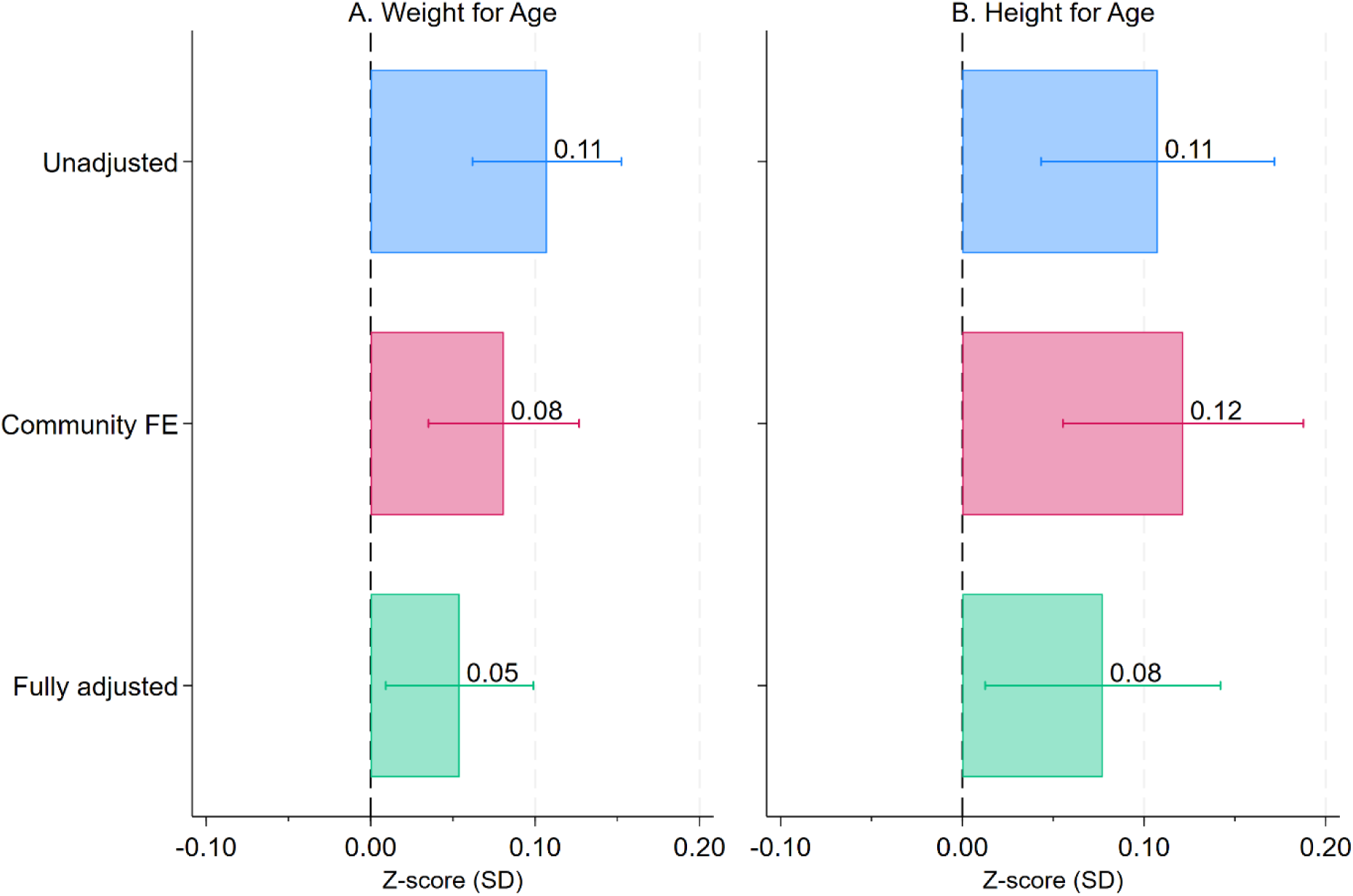
Triple difference coefficients on anthropometric outcomes comparing perinatal cash transfer beneficiaries to non-beneficiaries and firstborns to second born children between 2015 and 2021. Notes: Coefficients estimated using equation 1 (TD) are shown. Bands represent 95% confidence intervals. Community fixed effect model is a partially adjusted model which is stratified on community and includes community fixed effects. Covariates in the fully adjusted model include health insurance, family size, Hindu, Muslim, Christian, scheduled caste, scheduled tribe, socio-economic status score, mother’s height, mother’s age, mother’s education, child age, child sex, and COVID-19 lockdown. Standard errors are clustered at the community level.

Visualizations for pre-intervention trends in WAZ and HAZ across birth cohorts are shown in **Figure S3.** The figures show that parallel trends existed for WAZ and HAZ across CCT and non-CCT groups for first and second born children before 2017. However, since these visualizations do not directly test for parallel trends in a TD framework, we present the placebo regression estimates in **Table S4** to indirectly test the assumption. The placebo TD estimate is not statistically significant for WAZ and HAZ (p>0.10). Therefore, we do not reject the assumption of parallel trends for our main outcomes. Furthermore, as a robustness check for our main model, we ran the randomization inferences procedure (Heß, 2017) on the key outcome variables. **Table S5** shows that the triple interaction term’s coefficient in our primary fully adjusted model differs significantly from coefficients obtained from 1,000 random treatment assignments (p<0.05). In other words, the probability that our main results were obtained purely by chance is very low.

### 5.3 Potential pathways

We examine potential pathways to impacts on HAZ and WAZ by running regressions using the conditionalities of the PMMVY and evidenced based intermediate outcomes from systematic reviews (Manley et al., 2022). Conditionality related outcomes include pregnancy registration (binary), if the mother received four or more antenatal care (ANC) visits during pregnancy (binary), and whether the child received the full set of recommended immunizations (binary). Evidenced based outcomes included child level consumption of animal source foods (binary), whether the child achieved minimum diet diversity (binary), and diarrhoea incidence among children in the 15 days prior to the survey (binary) (Manley et al., 2022). Globally, the prevailing thesis suggest that poor households tend to spend more money on animal source foods compared to grains or tubers when they receive more money. We run these analyses with *equation 1* using a logit model for binary outcomes using the GEE GLM procedure described earlier.

Logit regressions on intermediate outcomes or pathways for impact (Figure 6) suggest that PMMVY works through its conditionalities: higher pregnancy registration (Odds ratio=1.12, p<=0.10), 4+ ANC (OR=1.06, p<0.05), and immunization (OR=1.10, p<0.001) among firstborn CCT beneficiaries in 2020 compared to the counterfactual. Earlier maternal CCTs in India, such as JSY, IGMSY, and Mamata reported similar results for vaccination uptake and antenatal checkups (Aizawa, 2022; De and Timilsina, 2020; Debnath, 2021; Lim et al., 2010; von Haaren and Klonner, 2021). For example, the Mamata scheme is estimated to have increased the odds of full immunization by 35% (Chakrabarti et al., 2021).

**Figure 6.**
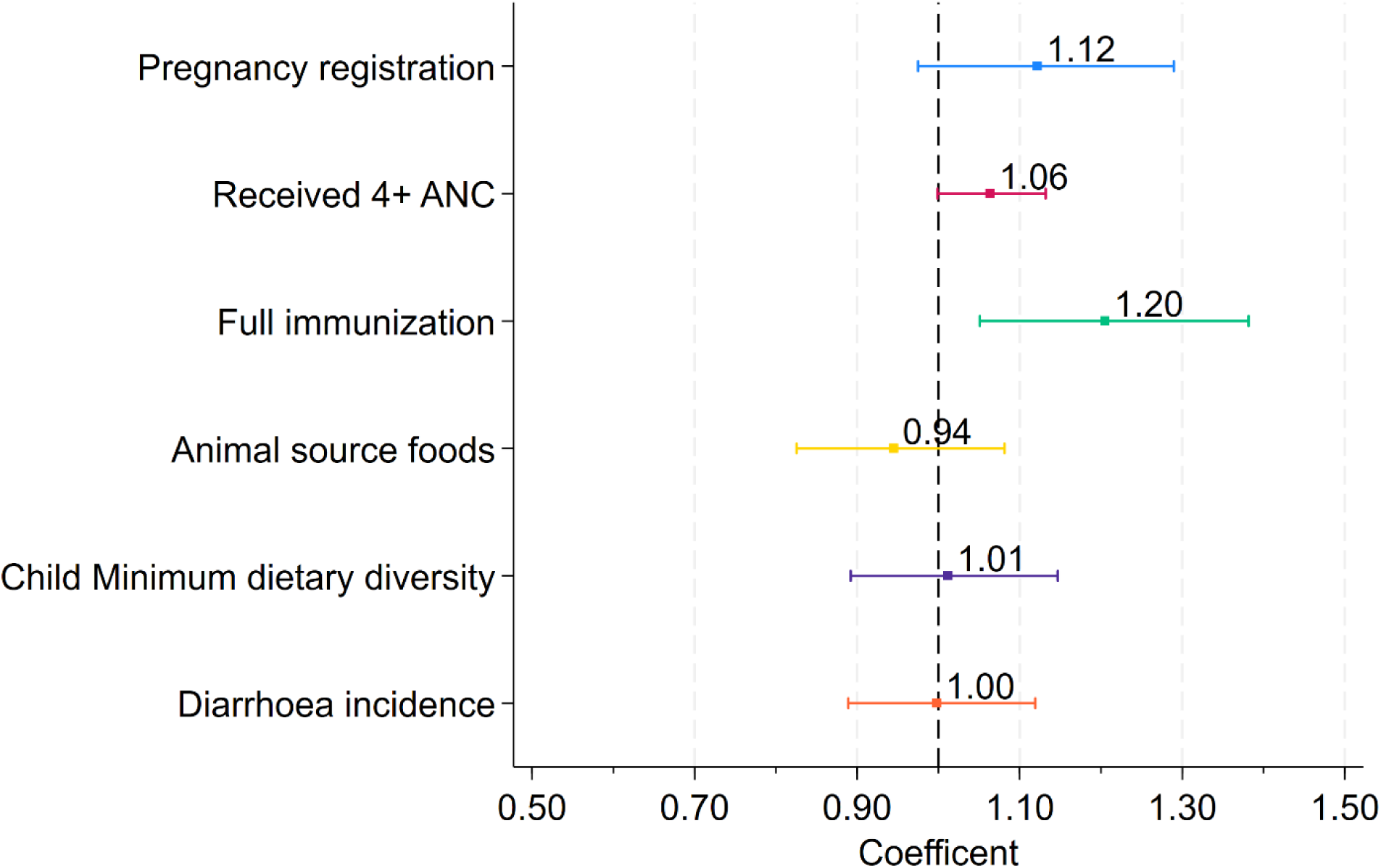
Pathways to impact: adjusted triple difference logistic regression coefficients of intermediate outcomes for PMMVY. Notes: All the outcome variables are in binary. Coefficients are from a logit regression Generalized Estimating Equation (GEE) with Generalized Linear Models (GLM) fit using equation 1 and an exchangeable or compound symmetry correlations structure on the community. Covariates in the fully adjusted logit model include health insurance, family size, Hindu, Muslim, Christian, scheduled caste, scheduled tribe, socio-economic status score, mother’s height, mother’s age, mother’s education, child age, child sex, and COVID-19 lockdown. The regressions were run using GEE with GLM fit is using equation 1 and an exchangeable or compound symmetry correlations structure on the community. The exchangeable correlation is similar to community fixed effects but more efficiently corrects point estimates assuming the correlation structure within each community is uniform across all pairs of observations within that community.

For other intermediate outcomes, there was no significant impact of PMMVY. A possible explanation for a lack of impact observed for diarrhoeal disease is that diarrhoea is reported as an incidence measure with a recall of 15 days in the NFHS. Given that diarrhoea is highly seasonal and the NFHS doesn’t visit households across all seasons, the outcome may not provide an accurate representation of diarrhoea prevalence. For dietary outcomes, a lack of impact on animal source foods may be due to large prevalence of vegetarianism in India (Ferry, 2023; Petrikova, 2022). Moreover, since the DHS uses a 24-hour dietary recall, for children older than 6 months, the survey timing likely does not align well with the window of the tranches. This may result in an underestimation of impacts on diet.

## 6 Heterogeneity in the Effects of the PMMVY Program

Large publicly funded cash transfer programs have a differential impact on beneficiary groups depending on contextual factors (Galiani and McEwan, 2013; Millán et al., 2019). In the Indian context, literature has demonstrated heterogeneous impacts of JSY based on income levels and awareness about government health programs (Debnath, 2021). Similarly, for the Mamata scheme in Odisha, a differential effect of the program was observed across child sex and household wealth (Kekre and Mahajan, 2023). Importantly, child sex was also differently distributed across first and second born children, and CCT groups and therefore could influence our TD results. Thus, we examine heterogeneity across Indian states via departmentalism, household wealth, and child sex.

### 6.1 Departmentalism

The PMMVY program provides flexibility for states and union territories to designate either the Ministry of Women and Child Development (MWCD), Ministry of Health and Family Welfare (MHFW), or Social Welfare and Justice (SWJ) departments for the program’s implementation, while the MWCD assumes the role of the national coordinating agency. Each of these departments possesses distinct mandates, organizational structures, and operational protocols, with the MWCD primarily responsible for coordinating initiatives related to women and children. Successful execution of a program of such magnitude necessitates sufficient human resources, expertise in managing social welfare schemes, and robust local infrastructure.

**Table S6** delineates the states that have opted for implementation through various ministries, including health, MWCD, and SWJ. We employed the WAZ and HAZ TD model specification across the three subgroups (Figure 7). In the adjusted models both WAZ (0.13 SD, p<0.001) and HAZ (0.19 SD, p<0.001) were significant for states where MHFW is responsible for implementing PMMVY. In the states where MWCD and SJW implements PMMVY, the results were modest and not significant. While it would be expected that the presence of departmental synergies at both national and state levels in MWCD should result in efficiency gains, our seemingly counterintuitive result is likely explained by the fact MHFW is more experienced at delivering national CCTs via JSY and may have achieved greater fidelity in program implementation through this mechanism.

**Figure 7.**
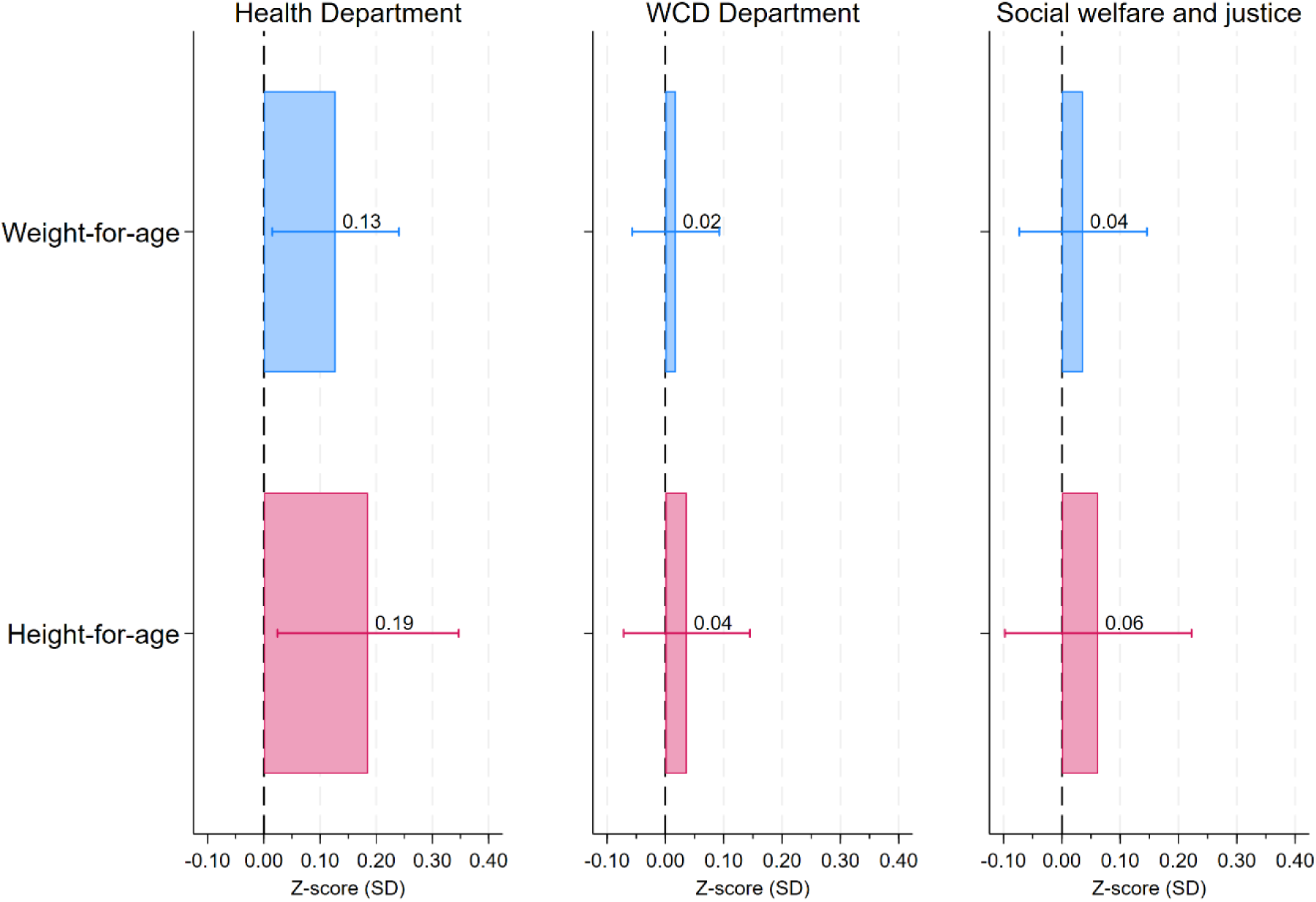
State level PMMVY implementation department’s heterogeneity effect on anthropometric outcomes using fully adjusted model. Note: Coefficients estimated using equation 1 (TD) and subsample are shown. Bands represent 95% confidence intervals. Covariates in the fully adjusted model include health insurance, family size, Hindu, Muslim, Christian, scheduled caste, scheduled tribe, socio-economic status score, mother’s height, mother’s age, mother’s education, child age, child sex, and COVID-19 lockdown. Standard errors are clustered at the community level.

### 6.2 Impact on the poor

CCTs programs like PMMVY are targeted towards women and children from poor households. Evidence from other Indian transfers have also shown larger benefits among the poor. For example, India’s flagship food subsidy programs, the Public Distribution System and Mid-day Meal Scheme, have documented pro-poor benefits (Chakrabarti et al., 2021; Kishore and Chakrabarti, 2015). Given the prior evidence, we expected PMMVY to also impact the poor differentially. Figure 8 shows larger coefficients on HAZ (0.10 SD, p<0.05) in poor sub-sample compared to the non-poor sub-sample (0.05 SD, p>0.1). The result on HAZ is likely attributable to the fact that mostly women from poor households access public health care in India and that ₹5000 is not likely to incentivize the non-poor to opt in. Moreover, this evidence also agrees with larger impacts on severely stunted children, who are also likely to belong to poor households.

**Figure 8.**
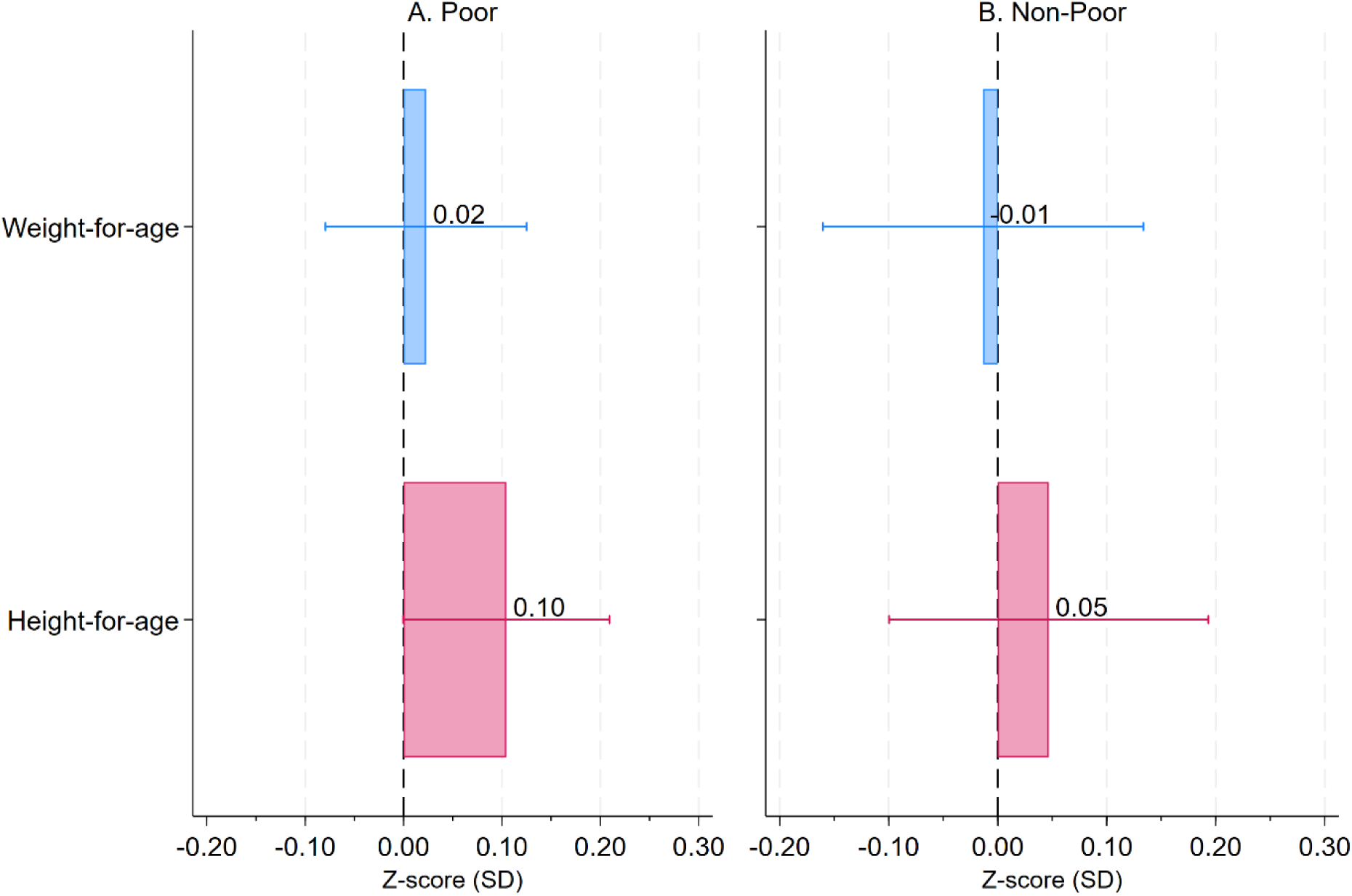
Heterogeneity effect on anthropometric outcomes based on household wealth levels using fully adjusted model. Note: Coefficients estimated using equation 1 (TD) and subsample are shown. Poor comprises of bottom three wealth quintiles while non-poor comprises of top two quintiles. Bands represent 95% confidence intervals. Covariates in the fully adjusted model include health insurance, family size, Hindu, Muslim, Christian, scheduled caste, scheduled tribe, socio-economic status score, mother’s height, mother’s age, mother’s education, child age, child sex, and COVID-19 lockdown. Standard errors are clustered at the community level.

Effects on WAZ were not significant in both groups. Coefficients for WAZ may have attenuated because these regressions were run on sub-samples of poor and non-poor. This statistically results in a different counterfactual for the TD model. For example, the TD in the poor sub-sample compares trends among poor CCT children to poor non-CCT children, whereas the TD in the full sample, includes poor and non-poor children.

### 6.3 Sex of the child

Gender-based discrimination and the preference for sons in India have been extensively documented and numerous studies have examined various aspects of this practice, including the prenatal determination of the child’s gender, postnatal neglect of the girl child’s dietary needs, and insufficient investment in the health and education of female children (Anukriti, 2018; Barcellos et al., 2014; Bharadwaj and Lakdawala, 2013; Borooah, 2004; Jayachandran, 2017; Jayachandran and Kuziemko, 2011). These factors may contribute to the heterogeneous impact of CCTs, whereby male children exhibit higher birth weights and WAZ compared to female children (Kekre and Mahajan, 2023).

Figure 9 demonstrates the varying effects of PMMVY on key anthropometric outcomes based on the sex of the child. Males (0.14 SD, p<0.05) have a higher and statistically significant impact on the PMMVY program compared to females (0.06 SD, p>0.1). One possible explanation for this disparity in anthropometric outcomes for girls is that female children have been reported to receive an inadequate diet, lower breastfeeding, vitamin supplementation and care after birth compared to male children (Barcellos et al., 2014; Borooah, 2004; Jayachandran and Kuziemko, 2011). It is likely that cash received from the program may have been more likely to be used to enhance growth and nutrition for males compared to females, especially if the male child was a first born. This is because of prevalent beliefs that first born males will eventually support the family financially or carry on the family legacy.

**Figure 9.**
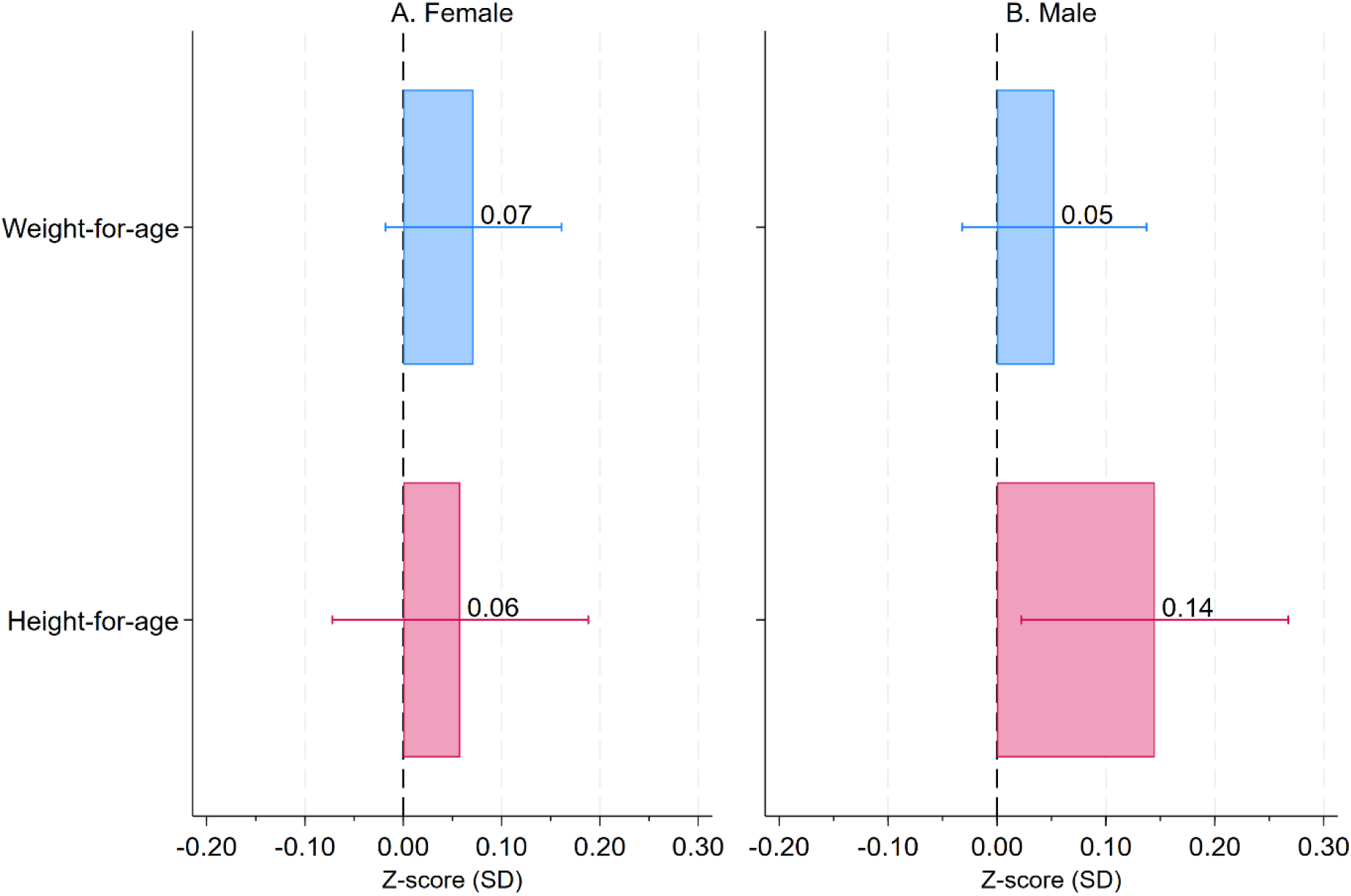
Heterogeneity effect on anthropometric outcomes based on child’s sex using fully adjusted model. Note: Coefficients estimated using equation 1 (TD) and subsample are shown. Bands represent 95% confidence intervals. Covariates in the fully adjusted model include health insurance, family size, Hindu, Muslim, Christian, scheduled caste, scheduled tribe, socio-economic status score, mother’s height, mother’s age, mother’s education, child age, child sex, and COVID-19 lockdown. Standard errors are clustered at the community level.

## 7 Economic analyses

### 7.1 Expenditure-outcome relationship

We use parliamentary expenditure data to estimate to the association between PMMVY spending at the state-year level and child anthropometry outcomes. For this, we restrict the sample to first born children and aggregate our data by state, birth-year, and CCT received (N=667). We fit a model with *equation 2* for state *s*, in birth-cohort *t*, CCT status *c*:

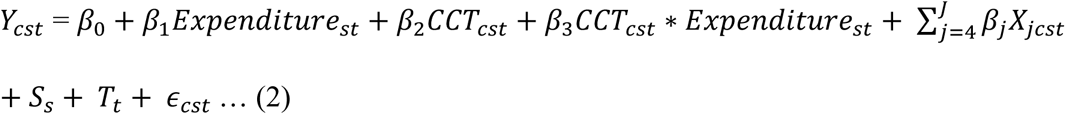

The model estimates the potential increase in HAZ, WAZ, stunting and underweight per amount spent on PMMVY. *Expenditure*_*st*_ varies by state and birth-cohort and represents an additional INR 1,000 per beneficiary spent on PMMVY. *CCT*_*cst*_ indicates whether the observation represents firstborn CCT beneficiaries in the state and birth cohort. The interaction, *β*_3_ estimates the dose response relationship between PMMVY money and undernutrition for firstborn CCT beneficiaries compared to firstborn non-CCT beneficiaries among firstborn children. The model adjusts for state fixed effects *S*_*s*_, birth year fixed effects *T*_*t*_, and the full set of household, mother, and child specific covariates *X*_*jcst*_.

There was heterogeneity in the coverage and expenditure on PMMVY across Indian states between 2017 and 2021 **(Table S7). Table 2** shows the effect of program expenditure on anthropometric outcomes using state-level birth cohort-specific per capita expenditure as a predictor. On average, for every additional INR 1,000 (approx.US$12) increase in the per capita spending on the eligible population, WAZ [0.063, p<0.001] and HAZ [0.066, p<0.01]) improved in the fully adjusted models that controlled for child, mother, household level covariates and fixed effects. Similarly, for every additional INR 1,000, underweight decreased by 1.27 percent points (p<0.05) and stunting by 1.38 percentage points (p<0.05), on average among firstborns.

**Table 2.** Association between anthropometric outcomes and state-level birth cohort-specific per capita expenditure on PMMVY among first-born children born between 2010 and 2021.

|  | **Partially**<br>**adjusted** | **Fully**<br>**adjusted** | **Partially**<br>**adjusted** | **Fully**<br>**adjusted** |
| --- | --- | --- | --- | --- |
|  | **Weight for Age (WAZ), SD** |  | **Height for Age (HAZ), SD** |  |
| PMMVY expenditure per-capita, 1000 Indian rupees | -0.034**<br>[-0.067, -0.001] | -0.033*<br>[-0.067, 0.000] | -0.052**<br>[-0.096, -0.009] | -0.059***<br>[-0.101, -0.017] |
| Perinatal cash beneficiary, proportion | -0.176***<br>[-0.237, -0.115] | -0.091**<br>[-0.174, -0.008] | -0.153***<br>[-0.223, -0.082] | -0.079<br>[-0.177, 0.018] |
| PMMVY expenditure * perinatal cash beneficiary | 0.081***<br>[0.043, 0.119] | 0.063***<br>[0.021, 0.105] | 0.079***<br>[0.026, 0.133] | 0.066**<br>[0.013, 0.118] |
|  | **Underweight (WAZ < -2 SD), %** |  | **Stunting (HAZ < -2 SD), %** |  |
| PMMVY expenditure per-capita, 1000 Indian rupees | 1.117**<br>[0.090, 2.145] | 1.118**<br>[0.188, 2.047] | 0.81<br>[-0.211, 1.830] | 0.878*<br>[-0.124, 1.880] |
| Perinatal cash beneficiary, proportion | 4.432***<br>[2.450, 6.414] | 1.702<br>[-0.490, 3.894] | 3.232***<br>[1.315, 5.149] | 1.257<br>[-1.048, 3.562] |
| PMMVY expenditure * perinatal cash beneficiary | -1.825***<br>[-3.002, -0.648] | -1.271**<br>[-2.468, -0.074] | -1.642***<br>[-2.871, -0.413] | -1.383**<br>[-2.613, -0.153] |
| Birth Year Controls | Yes | Yes | Yes | Yes |
| State Fixed Effects | Yes | Yes | Yes | Yes |
| Child Controls | No | Yes | No | Yes |
| Mother Controls | No | Yes | No | Yes |
| Household Control | No | Yes | No | Yes |
| N (state-birth year) | 648 | 648 | 648 | 648 |

Note: NFHS household data was aggregated by groups of state, birth year, birth order, and cash transfer received. PMMVY expenditure per capita is the estimated amount spent per eligible beneficiary in a specific state-birth year. Child, mother, and household controls include health insurance, family size, Hindu, Muslim, Christian, scheduled caste, scheduled tribe, socio-economic status score, mother's height, mother's age, mother's education, child age, and child sex. Confidence interval at 95% for the state level panel estimation are reported in the parentheses. \*\*\*, \*\*, \* show significance at 1%, 5% and 10%, respectively.

### 7.2 Benefit-cost analysis

Using *β*_3_ estimates from *equation 2* with underweight and stunting, we perform a benefit-cost analysis for PMMVY using economic analyses methods used in other studies (Chakrabarti et al., 2019). First, we multiply *β*_3_ by the mean values of *CCT*_*cst*_ in 2018, 2019 and 2020, respectively, to obtain the proportion of underweight and stunting prevalence averted by PMMVY in the population for those years. Second, we multiply the proportions obtained in step 1 by the total number of first-born children aged 0-1 years, who were underweight or stunted in the years 2018, 2019, and 2020 to obtain the total number of cases averted. Third, we multiply the underweight/stunting averted cases by their respective disability adjusted life year (DALY) rates. Fourth, we multiply the total DALYs by the per-capita gross domestic product (GDP) of India to obtain the total economic benefit of cases averted, and sum the benefit values for 2018, 2019 and 2020. Fifth, we divide the economic benefit by the total spending on the program in 2018, 2019 and 2020 to obtain the short run benefit-cost ratio for PMMVY.

While the primary objectives of PMMVY do not explicitly target stunting and underweight reduction, our study findings pertaining to HAZ and WAZ prompted an investigation into cases of stunting and underweight averted. Moreover, the availability of DALY data through the Global Burden of Disease study for underweight and stunting facilitated economic analyses. The research presented here contributes insights into the potential secondary benefits of PMMVY and its implications for decision making among policy makers.

Economic analyses showed that PMMVY delivered a combined health benefit of 47,064,813 DALYs everted due to underweight reduction and 26,759,834 DALYs averted due to stunting reduction **(Table 3)**. Assuming the economic value of a DALY is equal to per-head GDP, the combined economic benefit of PMMVY was US$1040 million for three years with a benefit-cost ratio of 1.35, suggesting that the program was cost effective in the short-run.

**Table 3.**
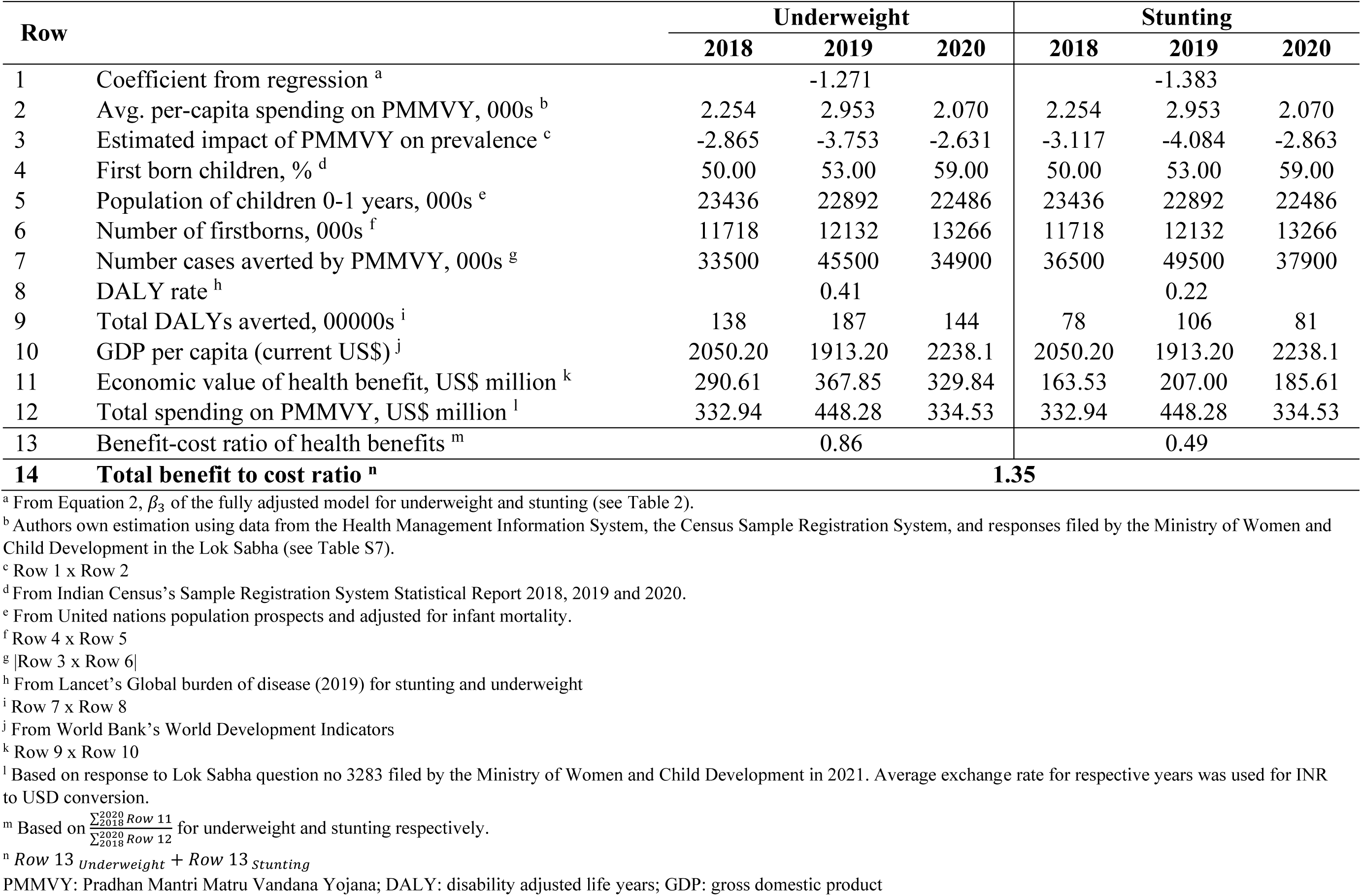
Benefit-cost analysis of the PMMVY between 2018 – 2020.

In previous research, it has been found that implementing essential interventions during the first 1000 days can result in a remarkable benefit-cost ratio of over 30 (Hoddinott et al., 2013). These estimates assume that the full range of interventions leads to a 20% reduction in undernutrition within the target population. However, our investigation reveals that despite its lower per-beneficiary cost, the PMMVY delivers a comparatively lower reduction in stunting when compared to targeted health interventions. Our calculated benefit-cost ratio of 1.35 focuses solely on immediate gains and does not account for the substantial long-term societal benefits arising from stunting reduction, which can result in even greater economic returns. Additionally, our analyses do not consider the health advantages associated with improved immunization and antenatal care coverage among program beneficiaries. Therefore, further exploration is needed to fully comprehend the potential impact of CCTs on child health and well-being in LMICs.

## 8 Discussion

### 8.1 Strengths and limitations

Strengths of our study include the use of nationally representative, repeated cross-sectional survey datasets that establish temporal order in that exposure to the PMMVY precedes the studied outcomes. We employed two sets of counterfactuals via our TD models that did not reject the assumption of parallel trends across both outcomes, and accounted for a comprehensive set of plausible confounders, thus supporting causal inference. Our study further provides insights in program pathways to impact, heterogeneity of coefficients via mechanisms like son preference, and estimates the economic viability of the program. To our knowledge, this is the first study that evaluates the PMMVY program with respect to nutrition outcomes among children in India. Our results are relevant for LMICs where perinatal CCTs are being implemented.

Our study is not without limitations. First, survey datasets used in our study do not provide precise information on actual receipt of the PMMVY among women in India. Our analyses estimate population level average treatment effects that rely on aggregate trends (Lachin, 2000). Second, owing to the nonrandomized nature of PMMVY receipt, confounding from unobserved variables that correspond with both exposure to PMMVY and child nutrition outcomes cannot be ruled out. Given our TD framework that comprehensively accounts for large potential confounders, any remaining biases could stem from unobserved factors that would 1) only affect firstborns but not second children within the CCT groups; 2) exhibit significant correlation with child anthropometry; and 3) not be completely accounted for by the control variables included in our analyses. Third, given the five-year gap between the two NFHS waves used in this study, maturation bias stemming from time-varying factors may likely attenuate our regression estimates (Handley et al., 2018). Fourth, our benefit-cost estimates assume that the value of each DALY averted is equal to per head GDP, however, valuing human suffering in monetary terms is challenging (Chakrabarti et al., 2019).

### 8.2 Barriers to implementation

A recent study commissioned in three states identified various hurdles that the beneficiaries face while availing the benefits of PMMVY (Sekher and Alagarajan, 2019). It highlighted the procurement of documentation for enrolling into PMMVY as the primary hurdle (Sekher and Alagarajan, 2019). **Table S2** highlights payment tranches, as well as the conditions and documentation linked with each installment. To be eligible for these benefits, women must fill out lengthy documents for each of the three instalments. Aside from connecting their bank account with Aadhaar (the Government of India’s individual identification system), they must also show their “mother-child protection” (MCP) card, husband’s and their own Aadhaar card, and bank passbook (Sekher and Alagarajan, 2019). Apart from these, cooperation of Anganwadi workers and the child development project officers is also necessary for successfully filling the online application (Dreze et al., 2021). These hurdles make the procedure cumbersome, especially for women with minimal education (Kalra and Priya, 2020). Furthermore, banking-related issues such as inaccessibility of financial services, bank officials’ reluctance to provide zero balance accounts, and PAN card non-availability are also observed (Dreze et al., 2021; Kalra and Priya, 2020; Sekher and Alagarajan, 2019). Even if beneficiaries are able to overcome these issues, women are far less likely than males to utilize and access their bank accounts and are thereby systematically excluded from the possible benefits of state-initiated transfers (Sabherwal et al., 2019).

Payment disbursement under PMMVY has often been delayed which restricts pregnant women’s ability to use the funds. It was also observed that the first instalment was often not provided to pregnant women in a timely manner. In some cases, beneficiaries got all three payments at once, after the childbirth (Sekher and Alagarajan, 2019). This subsequently leads to beneficiaries spending the cash transfer amount on covering regular household expenditure rather than nutritious foods at critical points in first 1000 days (Sekher and Alagarajan, 2019). **Table S7**, based on PMMVY data on disbursements and beneficiary reach, suggests that implementation varies greatly across states. This was also observed for the JSY, which was implemented nationally (Lim et al., 2010).

Despite these implementation challenges, our study finds that the program has a positive impact on child anthropometry outcomes, particularly HAZ and WAZ for those who received the PMMVY cash benefit. Further, our assessment shows a decrease in underweight and stunting on average over the first three years after PMMVY rollout. A benefit-cost analysis using the reduction shows that PMMVY produces a 35% return on the amount spent during that period.

### 8.3 Conclusion

Our study assessed the impact of the PMMVY, a national CCT program targeting pregnant and lactating women, on children’s nutrition outcomes, focusing on firstborns likely to benefit from the program. Analyzing multiple national surveys, we report that PMMVY is operational at-scale throughout India. However, despite its rights-based framework, the program presently fails to reach all entitled beneficiaries, primarily due to several implementation challenges. Nevertheless, our findings suggest that the program is associated with modest improvements in child anthropometric outcomes in a cost-effective manner, underscoring its potential to positively influence child health in India. With the extension of PMMVY to second-born girls in 2022, the program stands poised to cover millions more, emphasizing the need to address known barriers to access such as lengthy documentation and delayed payment disbursement.

## Data Availability

All data produced in the present study are available upon reasonable request to the authors.

## Supplementary tables and figures

**Table S1.** Landscape of India’s perinatal conditional cash transfer programs, 1987-2021.

| Scheme | Period | Region | Amount (INR) | Program Conditionality | Limits |
| --- | --- | --- | --- | --- | --- |
| Muthu Lakshmi Reddy Maternal Benefit Scheme (MRMBS) | 1987 – present | Tamil Nadu – state | 18,000 splits into many tranches | Antenatal care (ANC), Iron Folic Acid (IFA) supplements, deworming, vaccinations | 2 live births |
| National Maternity Benefit Scheme (NMBS) | 1995 – 2005 | India – national | ₹500 | N/A | 2 live births<br>Woman should be from Below Poverty Line (BPL) cardholder family |
| Janani Suraksha Yojana (JSY) | 2005 – present | India – national | 1,000 (rural) 1,400 (urban) | Institutional delivery | Institutional delivery in government hospital, BPL card holder/ Scheduled Tribes (ST) / Scheduled Castes (SC) in private hospital |
| Mamata scheme (O) | 2011 – present | Odisha – state | 5,000 splits into 4 tranches | Pregnancy registered, ANC, IFA, Nutrition counseling, Vitamin A supplements, vaccinations | First 2 live births |
| Indira Gandhi Matritva Suraksha Yojana (IGMSY) | 2011 – 2016 | India – national<sup>1</sup> | 4,000 splits into 3 tranches | Pregnancy registered, ANC, IFA, Nutrition counseling, first dose of child vaccinations (DPT, BCG, Polio) | First 2 live births |
| Pradhan Mantri Matru Vandana Yojana (PMMVY) | 2017 – present | India – national | 5,000 splits into 3 tranches<sup>2</sup> | Pregnancy registered, ANC, first dose of child vaccinations (DPT, BCG, Polio) | First live birth<sup>3</sup> |
| KCR Kit | 2017 – present | Telangana – state | 12,000 (Boy)<br>13,000 (Girl)<br>into 5 tranches | ANC, Institutional delivery, first and second dose of child vaccinations | First 2 live births, treatment in government hospital |
| Mamata scheme (G) | 2018 – present | Goa - state | 10,000 in 1 tranche | Institutional delivery | Live birth of girl child |

Note: Program details reflect the most recent identified for the scheme except in the case of PMMVY which is being evaluated in this paper. <sup>1</sup> Pilot program implemented in 52 out of the 640 districts in India. <sup>2</sup> At the time of writing under the new program guidelines released in March 2022, the number of instalments under the scheme has been reduced from 3 to 2 instalments. <sup>3</sup> As per the March 2022 guidelines, PMMVY benefits are being extended to a second child, but only if the second child is a girl. DPT stands for Diphtheria Pertussis & Tetanus vaccine and BCG stands for Bacillus Calmette Guerin single dose vaccine.

**Table S2.** Instalment wise PMMVY layout.

| **Instalment** | **Conditionality** | **Documents Required** | **Amount (INR)** |
| --- | --- | --- | --- |
| First Instalment (Registration) | • Register her pregnancy at any field functionary center along with required documents.<br>• Register her pregnancy within 150 days | • Application Form 1-A<br>• MCP Card<br>• Identity proof<br>• Bank/Post Office Account Passbook | 1,000 |
| Second Instalment | • At least one Ante Natal Care Check Up<br>• Can be claimed post 180 days of pregnancy | • Application Form 1-B<br>• MCP Card | 2,000 |
| Third Instalment | • Childbirth is registered<br>• Child has received first cycle of immunizations of BCG, OPV, DPT and Hepatitis B<br>• Aadhaar is mandatory in all states except for J&K, Assam, Meghalaya | • Application Form 1-C<br>• MCP Card<br>• Aadhaar ID<br>• Birth Certificate | 2,000 |

Source: (Sekher and Alagarajan, 2019); DPT stands for Diphtheria Pertussis & Tetanus vaccine, BCG stands for Bacillus Calmette Guerin single dose vaccine and OPV refers to Oral polio vaccine. MCP card stands for Mother child protection card, while Aadhaar refers to unique identification cards provided by the government.

**Figure S1.**
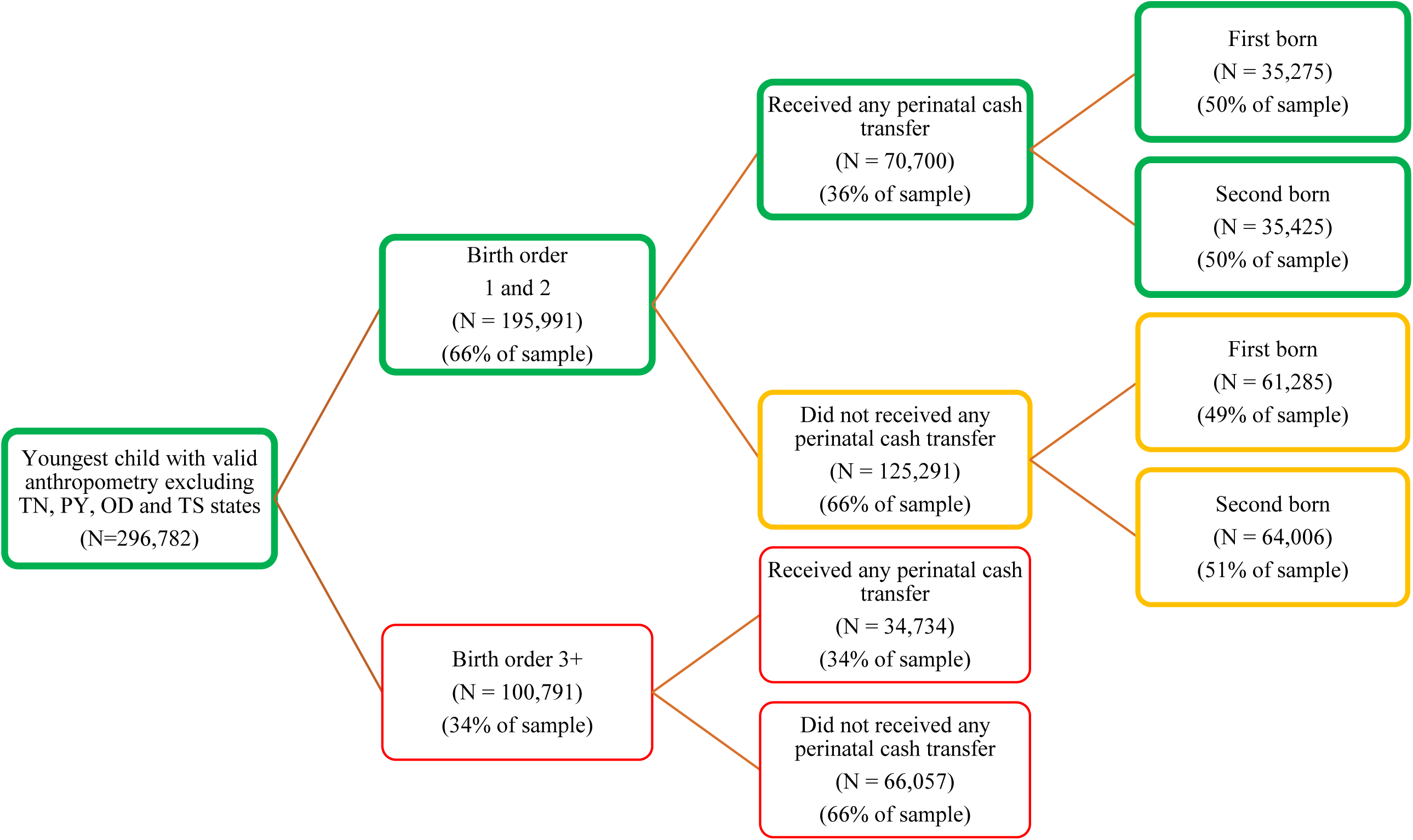
Sample restrictions and comparison groups used in the study.

**Figure S2.**
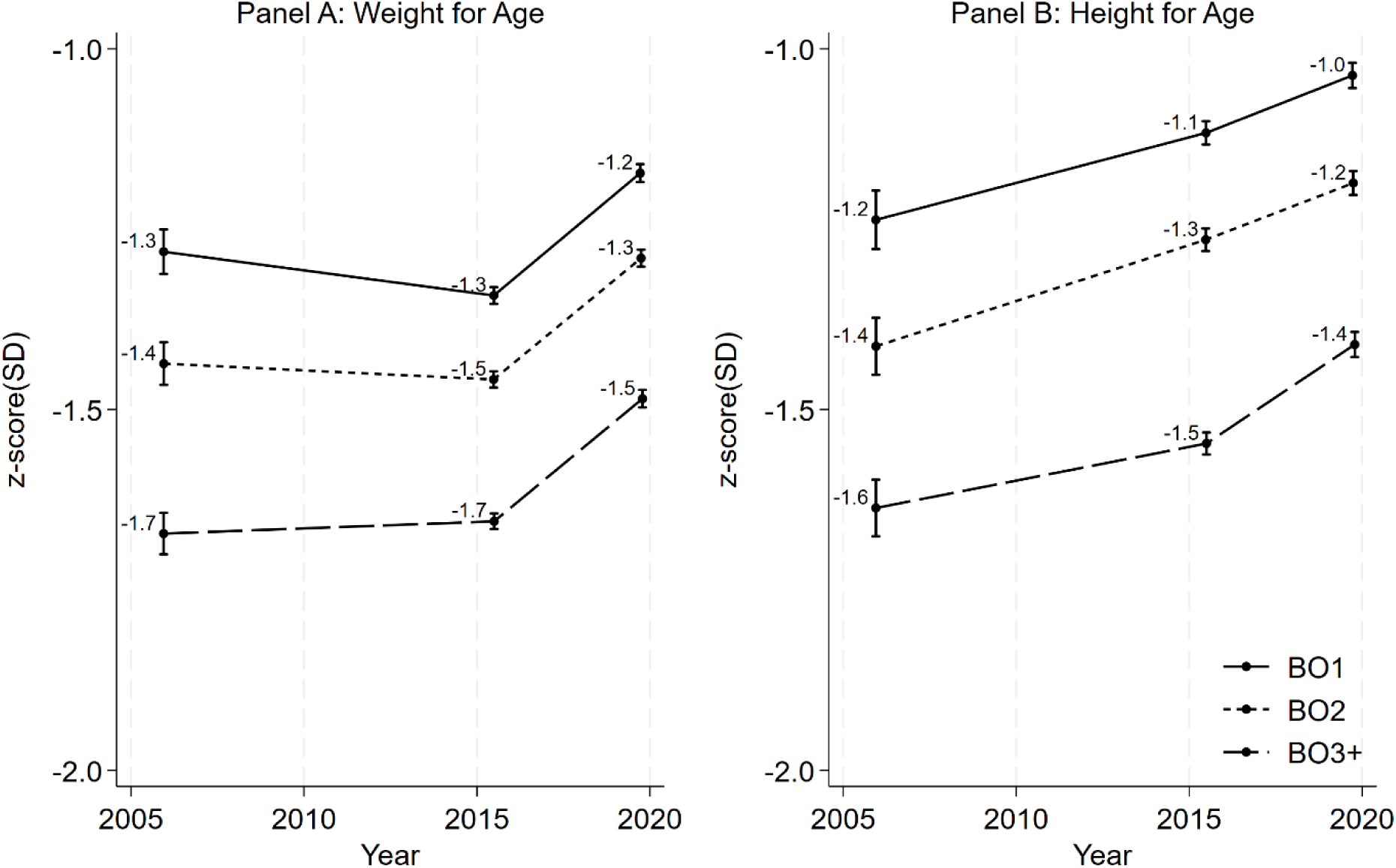
Trends in outcomes by birth order, 2005-2021. Notes: States with existing maternal benefits programs such as Tamil Nadu, Odisha, and Puducherry were dropped from the sample to isolate the PMMVY beneficiaries.

**Table S3.** Triple differences estimate of changes in anthropometric outcomes comparing perinatal cash transfer beneficiaries to non-beneficiaries and firstborns to second born children between 2015 and 2021.

|  | Weight for Age |  |  | Height for Age |  |  |
| --- | --- | --- | --- | --- | --- | --- |
|  | Unadjusted | Community fixed effects | Fully adjusted | Unadjusted | Community fixed effects | Fully adjusted |
| Non-CCT second-born children in 2016 ($\beta_0$) | -1.507***<br>[-1.518, -1.497] | -1.487***<br>[-1.496, -1.478] | -6.068***<br>[-6.218, -5.919] | -1.375***<br>[-1.389, -1.362] | -1.405***<br>[-1.418, -1.393] | -7.007***<br>[-7.218, -6.796] |
| CCT ($\beta_1$) | -0.099***<br>[-0.114, -0.083] | -0.050***<br>[-0.066, -0.035] | -0.024***<br>[-0.039, -0.009] | -0.139***<br>[-0.160, -0.119] | -0.104***<br>[-0.126, -0.082] | -0.067***<br>[-0.089, -0.046] |
| Firstborn ($\beta_2$) | 0.242***<br>[0.226, 0.258] | 0.153***<br>[0.136, 0.169] | 0.055***<br>[0.038, 0.072] | 0.326***<br>[0.304, 0.348] | 0.219***<br>[0.196, 0.242] | 0.068***<br>[0.045, 0.092] |
| Year (ref 2017) ($\beta_3$) | 0.222***<br>[0.204, 0.239] | 0.183***<br>[0.162, 0.203] | -0.109***<br>[-0.131, -0.086] | 0.214***<br>[0.189, 0.239] | 0.363***<br>[0.333, 0.393] | -0.366***<br>[-0.399, -0.333] |
| CCT * Firstborn ($\beta_4$) | -0.052***<br>[-0.078, -0.026] | -0.052***<br>[-0.079, -0.026] | -0.033**<br>[-0.059, -0.008] | -0.075***<br>[-0.110, -0.039] | -0.068***<br>[-0.105, -0.031] | -0.037**<br>[-0.073, -0.001] |
| CCT * Year ($\beta_5$) | -0.070**<br>[-0.097, -0.042] | -0.044**<br>[-0.072, -0.016] | -0.028**<br>[-0.056, 0.001] | -0.017<br>[-0.056, 0.022] | -0.044<br>[-0.085, 0.003] | 0.003<br>[-0.037, 0.043] |
| Firstborn * Year ($\beta_6$) | -0.073***<br>[-0.100, -0.045] | -0.046***<br>[-0.074, -0.018] | -0.014<br>[-0.042, 0.013] | -0.115***<br>[-0.155, -0.075] | -0.092***<br>[-0.133, -0.051] | -0.028<br>[-0.068, -0.013] |
| **CCT \* Firstborn \* Year ($\beta_7$)** | **0.107\*\*\***<br>**[0.062, 0.152]** | **0.081\*\*\***<br>**[0.035, 0.127]** | **0.054\*\***<br>**[0.009, 0.099]** | **0.108\*\*\***<br>**[0.043, 0.172]** | **0.122\*\*\***<br>**[0.055, 0.188]** | **0.077\*\***<br>**[0.013, 0.142]** |
| Fixed Effects | No | Yes | Yes | No | Yes | Yes |
| Individual covariates | No | No | Yes | No | No | Yes |
| Household covariates | No | No | Yes | No | No | Yes |
| **N** | **295,988** | **295,988** | **294,588** | **296,782** | **296,782** | **295,374** |

Notes: Coefficients estimated using equation 1 (TD) are shown. Covariates include urban residence, health insurance, family size, Hindu, Muslim, Christian, scheduled caste, scheduled tribe, socio-economic status score, mother's height, mother's age, mother's education, child age, child sex, COVID-19 lockdown and community fixed effects. Standard errors are clustered at the community level. Confidence interval at 95% are reported in the parentheses.
\*\*\*, \*\*, \* show significance at 1%, 5% and 10%, respectively

**Figure S3.**
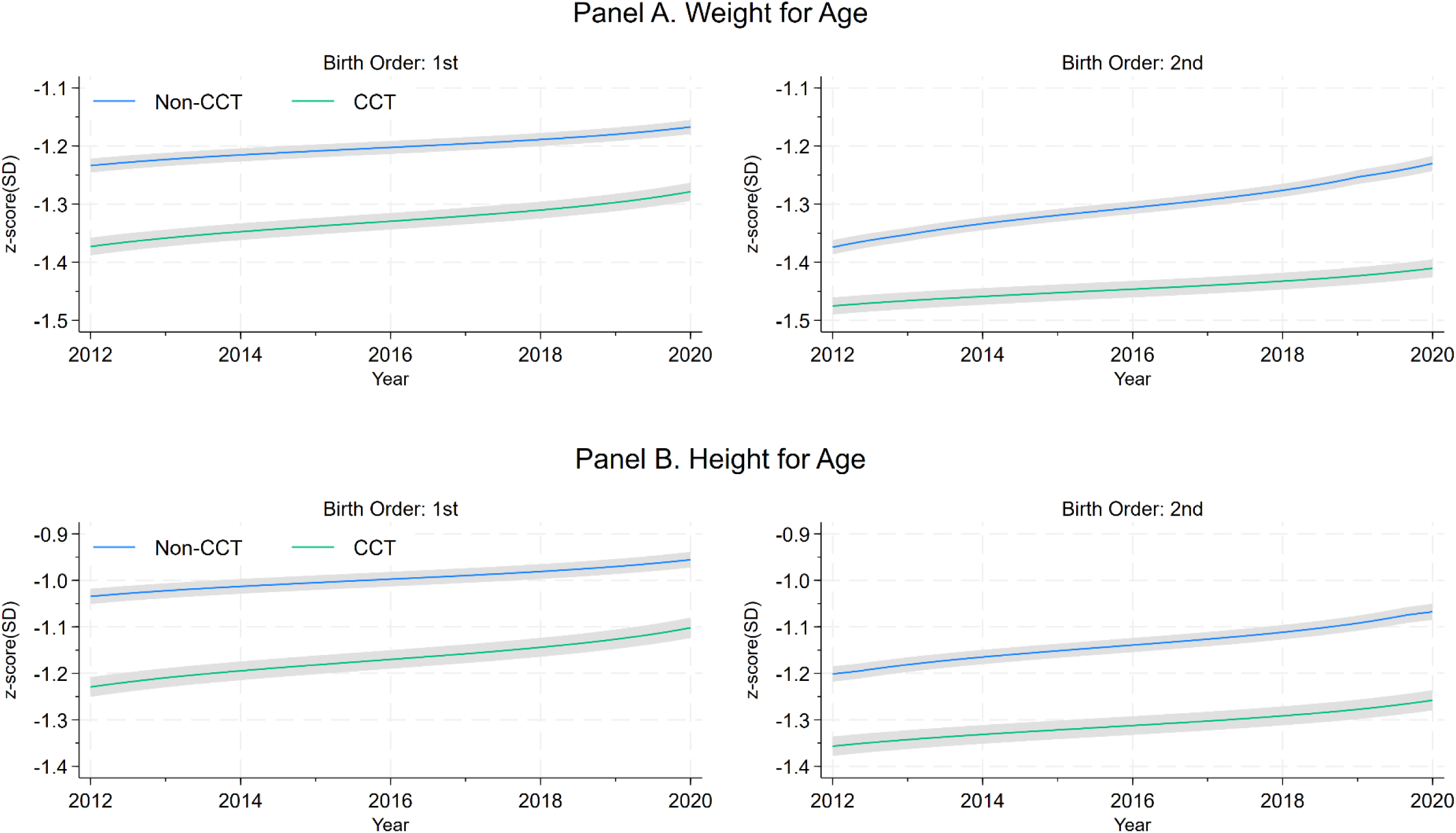
Pre-and-post intervention trends in child nutrition outcomes by cash transfer, birth order across birth cohorts. Notes: Plots represent the pooled data from NFHS 4 and 5. Weight for age and Height for age z-scores calculated between birth year 2012 and 2020 for birth order 1 (BO1) and birth order 2 (BO2), along with those who received conditional cash transfer (CCT) and those who didn’t (non-CCT). For parametric test of parallel trends see Table S4 in Supplementary material.

**Table S4.**
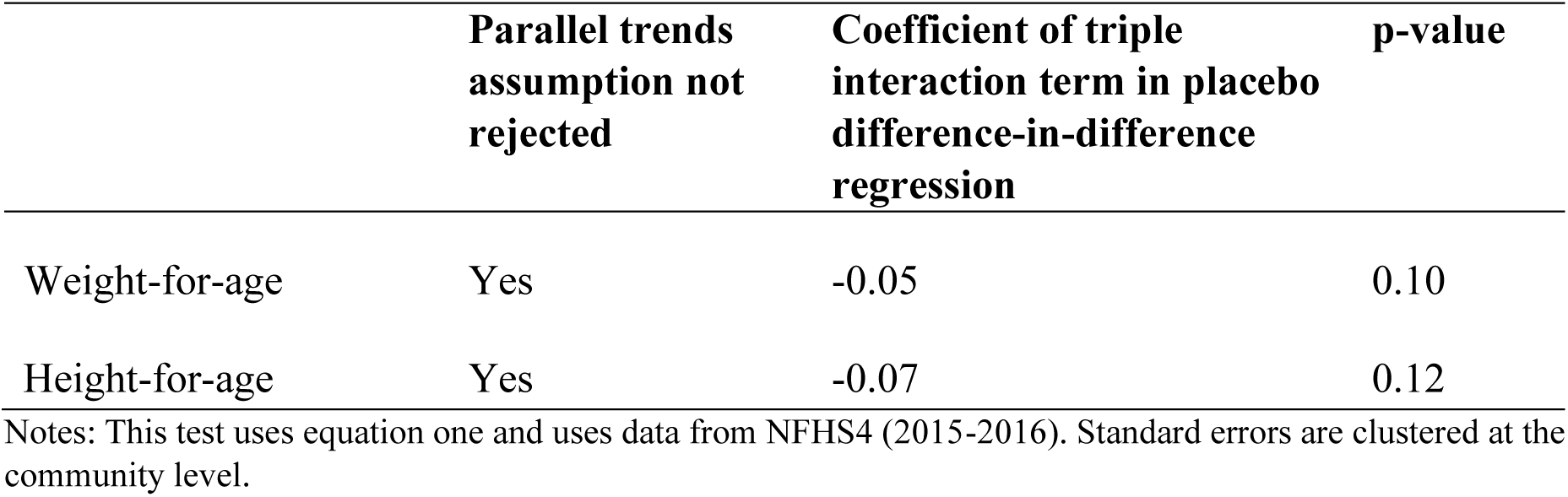
Test of parallel trends in preintervention period for the triple difference model (2010-2016)

|  | **Parallel trends**<br>**assumption not**<br>**rejected** | **Coefficient of triple**<br>**interaction term in placebo**<br>**difference-in-difference**<br>**regression** | **p-value** |
| --- | --- | --- | --- |
| Weight-for-age | Yes | -0.05 | 0.10 |
| Height-for-age | Yes | -0.07 | 0.12 |

Notes: This test uses equation one and uses data from NFHS4 (2015-2016). Standard errors are clustered at the community level.

**Table S5.** Randomization inference p-values on the triple differences estimate of changes in anthropometric outcomes.

|  | Unadjusted | Community fixed effect | Fully adjusted |
| --- | --- | --- | --- |
| Panel A: Weight for Age |  |  |  |
| Unadjusted p-value | 0.000 | 0.006 | 0.046 |
| RI adjusted p-value | 0.000 | 0.006 | 0.043 |
| Panel B: Height for Age |  |  |  |
| Unadjusted p-value | 0.052 | 0.005 | 0.043 |
| RI adjusted p-value | 0.051 | 0.003 | 0.036 |

Note: Unadjusted p-values are reported for the triple difference estimator shown in Table S3. The RI adjusted p-values are estimated using randomization inference procedure (Heß, 2017) using 1,000 random draws.

**Table S6.** Categorization of states based on the PMMVY implementation department.

| **Health Department** | **WCD Department** | **Social welfare and Justice (SWJ)** |
| --- | --- | --- |
| Andhra Pradesh | Arunachal Pradesh | Andaman & Nicobar |
| Chandigarh | Chhattisgarh | Assam |
| Dadra & Nagar Haveli | Delhi | Bihar |
| Daman & Diu | Goa | Himachal Pradesh |
| Meghalaya | Gujarat | Jammu & Kashmir (incl. Ladakh) |
| Telangana | Haryana | Kerala |
| Uttar Pradesh | Jharkhand | Manipur |
| West Bengal## | Karnataka | Mizoram |
|  | Lakshadweep | Nagaland |
|  | Madhya Pradesh | Sikkim |
|  | Maharashtra | Tripura |
|  | Punjab |  |
|  | Rajasthan |  |
|  | Uttarakhand |  |

Note: ## West Bengal didn't implement the program initially. But government notifications suggests that it is being implemented using Health department.

**Table S7.** State wise coverage and spending of PMMVY.

| States/Union Territories | Estimated % of first live born children PMMVY potentially covered |  |  |  | Estimated per capita spending on target population (In Indian rupees) |  |  |  |
| --- | --- | --- | --- | --- | --- | --- | --- | --- |
|  | 2017-18 | 2018-19 | 2019-20 | 2020-21 | 2017-18 | 2018-19 | 2019-20 | 2020-21 |
| Jammu & Kashmir | 29 | 42 | 40 | 47 | 96 | 1925 | 2004 | 1383 |
| Himachal Pradesh | 90 | 100 | 71 | 78 | 946 | 5845 | 4384 | 3720 |
| Punjab | 29 | 47 | 40 | 42 | 336 | 2381 | 2006 | 1659 |
| Chandigarh | 32 | 48 | 37 | 61 | 732 | 2043 | 1924 | 2733 |
| Uttarakhand | 44 | 77 | 71 | 83 | 661 | 3014 | 3676 | 3875 |
| Haryana | 40 | 75 | 44 | 44 | 548 | 3476 | 2511 | 1224 |
| Delhi | 22 | 40 | 31 | 48 | 256 | 1632 | 1640 | 1432 |
| Rajasthan | 19 | 100 | 43 | 34 | 105 | 3370 | 2297 | 1586 |
| Uttar Pradesh | 20 | 67 | 67 | 47 | 285 | 2337 | 3050 | 1866 |
| Bihar | 19 | 35 | 100 | 100 | 83 | 891 | 3982 | 6047 |
| Sikkim | 37 | 78 | 45 | 55 | 102 | 3377 | 3215 | 1761 |
| Arunachal Pradesh | 9 | 67 | 65 | 68 | 10 | 2076 | 2985 | 2754 |
| Nagaland | 2 | 32 | 100 | 75 | 0 | 1006 | 6613 | 3127 |
| Manipur | 23 | 37 | 100 | 89 | 324 | 1361 | 5016 | 3940 |
| Mizoram | 41 | 100 | 42 | 56 | 87 | 4813 | 3306 | 2532 |
| Tripura | 22 | 69 | 81 | 64 | 36 | 1797 | 4515 | 2259 |
| Meghalaya | 0 | 11 | 40 | 26 | 0 | 324 | 1613 | 944 |
| Assam | 9 | 45 | 97 | 53 | 34 | 1183 | 4756 | 2037 |
| West Bengal | 9 | 41 | 60 | 33 | 17 | 1446 | 2267 | 0 |
| Jharkhand | 33 | 45 | 49 | 38 | 213 | 1825 | 2289 | 1313 |
| Odisha | 0 | 0 | 0 | 0 | 0 | 0 | 0 | 0 |
| Chhattisgarh | 48 | 73 | 67 | 55 | 341 | 2456 | 3638 | 2449 |
| Madhya Pradesh | 80 | 100 | 88 | 96 | 587 | 5753 | 4876 | 4710 |
| Gujarat | 18 | 36 | 45 | 12 | 311 | 1819 | 2180 | 624 |
| Dadra & Nagar Haveli | 25 | 70 | 45 | 53 | 122 | 2214 | 2572 | 2095 |
| Maharashtra | 24 | 54 | 70 | 47 | 398 | 2148 | 3266 | 2158 |
| Andhra Pradesh | 63 | 85 | 61 | 54 | 876 | 4102 | 3942 | 1879 |
| Karnataka | 29 | 80 | 69 | 100 | 451 | 2939 | 3851 | 3356 |
| Goa | 27 | 49 | 32 | 40 | 502 | 2284 | 1573 | 1760 |
| Lakshadweep | 49 | 58 | 55 | 55 | 0 | 2427 | 1454 | 2399 |
| Kerala | 52 | 78 | 79 | 88 | 698 | 3379 | 4291 | 3201 |
| Tamil Nadu | 0 | 42 | 75 | 77 | 0 | 759 | 2580 | 2296 |
| Puducherry | 9 | 31 | 26 | 34 | 57 | 1266 | 1519 | 1210 |
| Andaman & Nicobar Islands | 83 | 62 | 43 | 55 | 1461 | 3542 | 3320 | 2887 |
| Telangana | 0 | 0 | 0 | 0 | 0 | 0 | 0 | 0 |
| **India** | **26** | **59** | **63** | **54** | **283** | **2254** | **2953** | **2070** |

Note: Author's own estimation using the data from the Health Management Information System, the Census Sample Registration System, and responses filed by the Ministry of Women and Child Development in the Lok Sabha.

